# Baseline associations between household air pollution exposure and blood pressure among pregnant women in the Household Air Pollution Intervention Network (HAPIN) multi-country randomized controlled trial

**DOI:** 10.1101/2023.01.23.23284847

**Authors:** Wenlu Ye, Ajay Pillarisetti, Oscar de León, Kyle Steenland, Jennifer L. Peel, Maggie L. Clark, William Checkley, Lindsay J. Underhill, Ashlinn Quinn, Kalpana Balakrishnan, Sarada S. Garg, John P. McCracken, Lisa M. Thompson, Anaité Díaz-Artiga, Ghislaine Rosa, Victor G. Davila-Roman, Lisa de las Fuentes, Aris T. Papageorghiou, Yunyun Chen, Jiantong Wang, F. c Thomas, the Household Air Pollution Intervention Network (HAPIN) trial Investigators

**Affiliations:** Gangarosa Department of Environmental Health, Rollins School of Public Health, Emory University, Atlanta, Georgia, USA; Environmental Health Sciences, School of Public Health, University of California, Berkeley, California, USA; Department of Environmental and Radiological Health Sciences, Colorado State University, Fort Collins, Colorado, USA; Division of Pulmonary and Critical Care, School of Medicine, Johns Hopkins University, Baltimore, Maryland, USA; Center for Global Non-Communicable Disease Research and Training, School of Medicine, Johns Hopkins University, Baltimore, Maryland, USA; Cardiovascular Division, Washington University School of Medicine, St. Louis, MO, USA; Berkeley Air Monitoring Group, Berkeley, California, USA; Department of Environmental Health Engineering, Sri Ramachandra Institute for Higher Education and Research (Deemed University), Chennai, India; Global Health Institute, Collage of Public Health, University of Georgia, Athens, Georgia, USA; Nell Hodgson Woodruff School of Nursing, Emory University, Atlanta, GA, USA; Center for Health Studies, Universidad del Valle de Guatemala, Guatemala; Department of Infectious and Tropical Diseases, London School of Hygiene & Tropical Medicine, London, UK; Nuffield Department of Women’s and Reproductive Health, University of Oxford, Oxford, UK; Department of Biostatistics and Bioinformatics, Rollins School of Public Health, Emory University, Atlanta, Georgia, USA

**Keywords:** Household Air Pollution, Solid Fuels, Blood Pressure, Pregnant Women

## Abstract

Cooking and heating using solid fuels can result in dangerous levels of exposure to household air pollution (HAP). HAPIN is an ongoing randomized controlled trial assessing the impact of a liquified petroleum gas stove and fuel intervention on HAP exposure and health in Guatemala, India, Peru, and Rwanda among households that rely primarily on solid cooking fuels. Given the potential impacts of HAP exposure on cardiovascular outcomes during pregnancy, we seek to characterize the relationship between personal exposures to HAP and blood pressure among pregnant women at baseline (prior to intervention) in the study. We assessed associations between PM_2.5_ (particulate matter with an aerodynamic diameter ≤2.5 μm), BC (black carbon), and CO (carbon monoxide) exposures and blood pressure at baseline, prior to intervention, among 3195 pregnant women between 9 and 19 weeks of gestation. We measured 24-hour personal exposure to PM_2.5_/BC/CO and gestational blood pressure. Multivariable linear regression models were used to evaluate associations between personal exposures to three air pollutants and blood pressure parameters. Trial-wide, we found moderate increases in systolic blood pressure (SBP) and decreases in diastolic blood pressure (DBP) as exposure to PM_2.5_, BC, and CO increased. None of these associations, however, were significant at the 0.05 level. HAP exposure and blood pressure associations were inconsistent in direction and magnitude within each country. We observed effect modification by body mass index (BMI) in India and Peru. Compared to women with normal weights, obese women in India and Peru (but not in Rwanda or Guatemala) had higher SBP per unit increase in log transformed PM_2.5_ and BC exposures. We did not find a cross-sectional association between HAP exposure and blood pressure in pregnant women; however, HAP may be associated with higher blood pressure in pregnant women who are obese, but this increase was not consistent across settings.

## 1. INTRODUCTION

Approximately 49% of the global population – about 3.8 billion people – burn solid fuels (including coal, wood, charcoal, dung, and crop residues, among others) as an energy source for cooking and heating (1). These practices release high concentrations of air pollutants, resulting in exposures that often exceed WHO air pollution guidelines (2,3). Household air pollution (HAP) generated from the incomplete combustion of these fuels in traditional stoves contributes to a large burden of ill health – between 1.6 and 4 million deaths per year from causes including diabetes, respiratory, and cardiovascular diseases (CVD) (4,5).

Numerous pollutants are released during the combustion of solid fuels (6,7); the most well studied are (A) particulate matter with an aerodynamic diameter of 2.5 µm or less (PM_2.5_) and (B) carbon monoxide (CO) (8). As a potent short-lived climate pollutant and an important component of PM, black carbon (BC) also receives substantial attention, especially in recent household clean energy programs, given the co-benefits of reducing BC for both climate and health (9–11).

These pollutants have been associated with changes in blood pressure – a known risk factor for cardiovascular disease – in association with exposure to ambient air pollution (12) and among households using solid fuels (13–19). The exact mechanisms by which air pollution impacts blood pressure are unknown; it is hypothesized that pulmonary and systemic oxidative stress, inflammation, and disturbances of the cardiac autonomic nervous system may contribute to the observed effects (20–22).

While several studies have evaluated the relationship between solid fuel use and blood pressure, only a small number have measured personal exposure to HAP constituents. Others have focused on comparing biomass versus clean fuel users as a proxy for exposure (23–27). A number of studies have evaluated interventions that reduce HAP exposure and their impacts on blood pressure (16,18,28). However, few have evaluated the impact of HAP on blood pressure during pregnancy (29–32).

Blood pressure changes during pregnancy are well documented: during normal pregnancy, changes in blood volume and cardiac output result in decreased systolic and diastolic blood pressure in early pregnancy, followed by elevated blood pressure and a return to normal, pre-pregnancy levels (33–37). Elevated blood pressure complicates an estimate of 3-10% of pregnancies worldwide (38) and contributes to 30,000 maternal deaths annually (39).

The Household Air Pollution Intervention Network (HAPIN) multi-country (i.e., Guatemala, India, Peru, and Rwanda) randomized controlled trial (RCT) is evaluating the impact of a clean fuel and stove intervention during pregnancy on birth weight, growth, and severe pneumonia in children and blood pressure in older adult women over ∼18 months of follow-up. As secondary measures, HAPIN assessed, among other things, personal exposures of pregnant HAPIN participants to PM_2.5_, BC, and CO and their blood pressure at various time points throughout the trial (40,41).

Prior to randomization and intervention delivery, HAPIN collected baseline data on participants and their households and measured their systolic blood pressure (SBP) and diastolic blood pressure (DBP). We also assessed their personal exposures to household air pollutants. In this paper, we report 1) the trial-wide associations of HAP (i.e., PM_2.5_, BC, and CO) exposures and gestational blood pressure during this pre-intervention baseline period, and 2) country-specific associations in Guatemala, Peru, and Rwanda (results from India have been reported elsewhere (42)).

## 2. METHODS

### 2.1 Study Design, Location, and Population

This analysis includes 3195 pregnant women enrolled between 2016 and 2018 at the four predominantly rural international research centers (IRCs) of the HAPIN trial at baseline: Guatemala (N = 800), India (N = 799), Peru (N = 798), and Rwanda (N = 798). Details about the study design and locations are provided elsewhere (40) and briefly summarized below.

In Guatemala, the study sites are in the Jalapa municipality. Cooking occurs primarily indoors using wood in chimney stoves and open fires. In India, study sites are spread across two districts in the southern state of Tamil Nadu, where traditional clay/mud stoves are fueled with wood, predominantly indoors. In Peru, study sites are spread across six rural provinces in the Department of Puno. Households rely on dung-fueled stoves for daily cooking. The study sites in Rwanda are three neighboring sectors in the Kayonza District (Eastern Province). Most households cook indoors using traditional open stoves made of clay and bricks fueled with wood (*rondereza)* and portable charcoal stoves (*imbabura*).

Pregnant women were eligible for enrollment into the HAPIN trial if they 1) were between 18 and <35 years of age, 2) cooked primarily with solid fuels and did not plan to switch to clean fuels predominantly in the near future, 3) lived in the study area and did not plan to move permanently in the next 12 months, 4) were between 9 and < 20 weeks of gestation with a singleton pregnancy confirmed by ultrasound, 5) continued pregnancy at the time of randomization, 6) were not current smokers, and 7) agreed to participate with informed consent (40).

Study protocols and procedures have been reviewed and approved by institutional review boards (IRBs) or Ethics Committees of Emory University (00089799), Johns Hopkins University (00007403), Sri Ramachandra Institute of Higher Education and Research (IEC-N1/16/JUL/54/49) and the Indian Council of Medical Research – Health Ministry Screening Committee (5/8/4-30/(Env)/Indo-US/2016-NCD-I), Universidad del Valle de Guatemala (146-08-2016) and Guatemalan Ministry of Health National Ethics Committee (11-2016), Asociación Beneficia PRISMA (CE2981.17), the London School of Hygiene and Tropical Medicine (11664-3), the Rwandan National Ethics Committee (No.016/RNEC/2018), and Washington University in St. Louis (201611159).

### 2.2 Measurement of Personal Exposure to Household Air Pollution

Exposure measurement procedures have been published elsewhere (41). 24-hour personal exposures to PM_2.5_ and CO were measured simultaneously for all participants; BC concentrations were assessed using transmissometry after gravimetric sample collection. Pregnant women wore an IRC-specific customized garment, with exposure instrumentation kept near the breathing zone. Participants were also asked to hang the garment on a stand and keep it nearby (within 1-2m) when sleeping, bathing, or conducting other activities during which it was not suitable to wear the monitoring equipment.

Personal exposure to PM_2.5_ was monitored using the Enhanced Children’s MicroPEM™ (ECM) (RTI International, Research Triangle Park, USA). The ECM is lightweight (∼150g) and generates minimal noise. It measures real-time PM_2.5_ concentrations at 10-second intervals using a nephelometer and simultaneously collects an integrated gravimetric sample on a 15mm PTFE filter. The instrument also records temperature, relative humidity, inlet pressure, and triaxial accelerometry. Twenty-four-hour gravimetric samples were collected for each participant; changes in pre- and post-sampling filter mass were assessed using 1-µg resolution microbalances (Sartorius Cubis, MSA6.6s-000-DF, Göttingen, Germany) at the University of Georgia (for samples collected in Guatemala, Peru, and Rwanda) and Sri Ramachandra Institute for Higher Education and Research (for samples collected in India). Detailed methods and validity criteria are reported elsewhere (43). When a gravimetric sample was deemed invalid, due to missing or damaged filters or flow faults, nephelometer data was used to estimate PM_2.5_ concentrations normalized to per-device field-based filters. Quality control and assurance, duplicates, and wearing compliance are described elsewhere (43).

24-hour BC concentrations were estimated from PM_2.5_ filter samples using SootScan Model OT21 Optical Transmissometers (Magee Scientific, USA). BC depositions were estimated per Garland et al. (2017) (44). Personal CO exposure was measured by Lascar EL-USB-300 (Lascar Electronics, USA). The Lascar is the size of a large pen (125 × 26.4 × 26.4mm, 42g) and logs CO concentrations at 1-minute intervals. Details for CO and BC data quality assurance and instrument calibration are reported in (43).

### 2.3 Measurement of Blood Pressure

Following the 24-hour exposure assessment period, a nurse or trained field worker measured resting blood pressure in the right arm of the pregnant women in triplicate (with at least 2 minutes between measurements) using an automatic monitor (model HEM-907XL; Omron^®^) at the participant’s home. Before starting the measurement, the participant was instructed to sit on a chair in a quiet room for 5 min with legs uncrossed, their back supported by the chair, and their arm supported on a table. The pregnant woman also confirmed that she had not smoked, consumed alcohol, or caffeinated beverages (coffee, tea, or Coca-Cola), or cooked using biomass in the past 30 minutes. If she had done any of those activities in the 30 minutes prior to the measurement, she would be asked to refrain from doing these activities for 30 minutes before proceeding with the measurements.

A participant with a measured SBP ≥ 140 mmHg and/or a DBP ≥ 90 mmHg was checked again during the same visit. If the same result was observed on two measurements, the participant was referred to the nearest health center or hospital to receive age-appropriate treatment. If a participant had SBP < 80 mmHg or a DBP < 40 mmHg, she would also be referred to the nearest health center or hospital. In analyses, the average of all three blood pressure measurements was used, as no “white-coat” effect was observed. There were no implausible high blood pressure values. SBP values less than 70 mmHg and DBP values less than 35 mmHg were excluded as implausible. At baseline, no blood pressure value was excluded based on above criteria.

### 2.4 Questionnaires and Other Measurements

Questionnaires were administered by trained field workers in the local language to obtain information on households’ demographic and socioeconomic status; stove and energy use patterns; kitchen configuration(s); other exposure sources (e.g., environmental tobacco smoke; incense and garbage burning, etc.); self-reported medical/gynecological history and medication use; and lifestyle behaviors (i.e., physical activity, diet diversity, food insecurity, and alcohol/tobacco consumption). Questionnaires were tested prior to implementation. Baseline maternal weights (seca 876/874 scales; Seca) and heights (seca 213 stadiometer; Seca) were measured in duplicate. Gestational age at the blood pressure measurement was calculated by using the ultrasound-estimated gestational age at screening plus the difference in days between the screening date and the blood pressure measurement date.

### 2.5 Statistical Analysis

We used univariable and multivariable regression models to investigate the association between personal exposure to PM_2.5_/BC/CO and gestational SBP and DBP. As secondary endpoints, we also examined the associations of HAP exposures with 1) pulse pressure (PP), defined as *SBP* − *DBP* and 2) mean arterial pressure (MAP), defined as *DBP* + (*SBP* − *DBP*)/3, given their relevance to adverse pregnancy outcomes (45,46). We evaluated correlations between pollutants (Spearman’s *ρ*). Given their moderate to high correlations (PM_2.5_ and BC: Spearman’s *ρ* = 0.79; PM_2.5_ and CO: Spearman’s *ρ* = 0.47; CO and BC: Spearman’s *ρ* = 0.42), we ran separate models for PM_2.5_, BC and CO. Model assumptions were verified using routine regression diagnostics.

To account for the heterogeneity in HAP exposure, gestational blood pressure, and other factors across IRCs – and to allow for country-specific estimates – we conducted separate assessments for the associations between HAP exposure and gestational blood pressure in Guatemala, Peru and Rwanda, following a procedure similar to one performed previously in India (42). We then estimated the trial-wide association by combining IRC-specific estimates using the DerSimonian and Laird random effects method (47). In cases that the heterogeneity statistic Q was less than the degrees of freedom (df = 3), a fixed effects combined measure, using the inverse variance method, was estimated.

Because we hypothesized that there may be nonlinear relationships between HAP exposure and gestational blood pressure, we utilized Generalized Additive Models (GAMs) with default thin plate regression splines with 2, 3, and 4 degrees of freedom and restricted maximum likelihood estimation to model smooth functions of exposure. Highly skewed exposure data also motivates the use of a log scale for exposure (log-linear). We compared GAMs and parametric (linear and log-linear) models through visual inspection and a comparison of Akaike Information Criterion (AIC; an estimate of goodness of fit; a lower AIC indicates a better fit to the data). Based on these assessments, we report our main results for PM_2.5_/BC/CO and SBP/DBP relationships using linear models with (1) natural log-transformed and (2) categorical (in quartiles) exposure terms.

Potential confounders were included in the model if they changed the estimate of HAP exposure by more than 10% in each IRC and were evaluated as non-colliders using a directed acyclic graph (DAG) (Figure S1). Maternal age, gestational age, and BMI are included in all models as known confounders. Variables evaluated as potential confounders included nulliparity (defined as zero pregnancies reaching 20 weeks and 0 days of gestation or beyond; miscarriages can have occurred in a woman who is nulliparous), mother’s highest level of education, physical activity, date (weekday vs. weekend) and time (morning vs. afternoon) of the blood pressure measurement, household food insecurity score, mother’s minimum diet diversity, exposure to environmental tobacco smoke, and month of the blood pressure measurement (to account for potential seasonality). Potential effect modification by gestational age at the blood pressure measurement (median), maternal age (median), BMI (underweight, normal weight, overweight, and obese), physical activity (quartiles), and exposure to environmental tobacco smoke (Yes/No) was assessed using multiplicative interaction terms between these factors and PM_2.5_/BC/CO exposure variables in adjusted models. Additionally, we evaluated whether our results changed after excluding the highest and lowest 2.5% (total 5%) of exposure measurements, as few data points were collected at these extreme values.

All statistical modeling was using R version 4.0.3. GAMs were fitted using the ‘mcgv’ package (48). Mixed effects models were fitted with the ‘lme4’ package (49).

## 3. RESULTS

### 3.1 Participant characteristics

Five pregnant women were taking antihypertensive medication at baseline, and they were excluded from all analyses; the remaining 3190 pregnant women comprise our analytical population (**Table 1**). The average maternal age of this cohort was 25.4 years (range 18 – 35), and the mean gestational age was 15.4 (range 9.0 – 24.9) weeks (**Figure S2**). More than half (59%) of the participants were normal/healthy weight. 30% of the participants were considered overweight or obese; they were mainly in Guatemala and Peru. Conversely, India accounted for nearly all underweight pregnant women (308 out of 333). 1228 (38%) of the pregnant women were nulliparous and the percentage of nulliparity was highest in India. 422 (13%) of the participants had a history of spontaneous abortion. Few (3%) participants reported a history of preterm birth and stillborn. Very few participants reported a history of hypertension (<1%), or diabetes (<1%) at the baseline health assessment. About two-third of the women (2150, 67%) had completed at least primary education, and more than half (57%) were employed outside the household. 82% of the pregnant women were the primary cooks of their family. 334 (10%) women reported that one or more smokers lived in their household; 253 were in India, followed by Guatemala (44) and Rwanda (30).

**Table 1.** Baseline characteristics of pregnant women participating in the HAPIN Trial^1^

| Characteristic | Guatemala<br>N = 800 | India<br>N = 799 | Peru<br>N = 798 | Rwanda<br>N = 793 | All<br>N = 3190 |
| --- | --- | --- | --- | --- | --- |
| **Maternal characteristics** |  |  |  |  |  |
| Maternal age, Mean (SD) [Range] | 24.7 (4.4)<br>[18.0, 34.9] | 24.0 (3.8)<br>[18.1, 34.8] | 25.5 (4.5)<br>[18.0, 35.0] | 27.3 (4.4)<br>[18.1, 34.9] | 25.4 (4.5)<br>[18.0, 35.0] |
| Gestational age (weeks), Mean (SD) [Range] | 14.3 (3.1)<br>[9.0, 21.3] | 16.0 (3.0)<br>[9.6, 24.9] | 15.7 (3.4)<br>[9.0, 22.9] | 15.5 (2.8)<br>[9.7, 21.7] | 15.4 (3.1)<br>[9.0, 24.9] |
| BMI, Mean (SD) [Range] | 23.8 (3.3)<br>[16.4, 44.2] | 19.7 (3.2)<br>[13.3, 37.6] | 26.0 (3.6)<br>[17.9, 39.6] | 23.4 (3.4)<br>[16.6, 42.7] | 23.2 (4.1)<br>[13.3, 44.2] |
| BMI categories, N (%) |  |  |  |  |  |
| Underweight (<18.5) | 11 (1%) | 308 (39%) | 1 (<1%) | 13 (2%) | 333 (10%) |
| Normal/Healthy Weight (18.5 – 24.9) | 543 (68%) | 434 (54%) | 330 (41%) | 573 (72%) | 1880 (59%) |
| Overweight (25.0 – 29.9) | 205 (26%) | 50 (6%) | 341 (43%) | 166 (21%) | 762 (24%) |
| Obesity (≥30.0) | 36 (5%) | 7 (1%) | 116 (15%) | 37 (5%) | 196 (6%) |
| Missing | 5 (<1%) | 0 | 10 (1%) | 4 (1%) | 19 (1%) |
| Nulliparous, N (%) |  |  |  |  |  |
| Yes | 227 (28%) | 459 (57%) | 310 (39%) | 232 (29%) | 1228 (38%) |
| No | 573 (72%) | 340 (43%) | 484 (61%) | 559 (70%) | 1956 (61%) |
| Missing | 0 | 0 | 4 (1%) | 2 (<1%) | 6 (<1%) |
| History of preterm birth, N (%) | 19 (2%) | 12 (2%) | 36 (5%) | 24 (3%) | 91 (3%) |
| History of spontaneous abortion, N (%) | 123 (15%) | 84 (11%) | 84 (11%) | 131 (17%) | 422 (13%) |
| History of stillborn, N (%) | 35 (4%) | 15 (2%) | 13 (2%) | 37 (5%) | 100 (3%) |
| History of hypertension, N (%) | 7 (1%) | 0 | 3 (<1%) | 5 (<1%) | 15 (<1%) |
| History of diabetes, N (%) | 1 (<1%) | 0 | 1 (<1%) | 3 (<1%) | 5 (<1%) |
| Mother's education level, N (%) |  |  |  |  |  |
| No formal education or Primary school incomplete | 381 (48%) | 285 (36%) | 35 (4%) | 338 (43%) | 1039 (33%) |
| Primary school complete or Secondary school incomplete | 312 (39%) | 227 (28%) | 234 (29%) | 316 (40%) | 1089 (34%) |
| Secondary school complete or Vocational or Some college or university | 107 (13%) | 287 (36%) | 528 (66%) | 139 (18%) | 1061 (33%) |
| Missing | 0 | 0 | 1 (1%) | 0 | 1 (<1%) |
| Mother's occupation, N (%) |  |  |  |  |  |
| Agriculture | 6 (1%) | 338 (42%) | 513 (64%) | 540 (68%) | 1397 (44%) |
| Commercial | 17 (2%) | 4 (1%) | 111 (14%) | 135 (17%) | 267 (8%) |
| Household | 746 (93%) | 432 (54%) | 132 (17%) | 56 (7%) | 1366 (43%) |
| Other | 31 (4%) | 25 (3%) | 42 (5%) | 62 (8%) | 160 (5%) |
| Physical activities (total MET-min/day), Mean (SD) |  |  |  |  |  |
| Quartile 1 | 11.8 (9.6) | 109 (48.1) | 646 (269) | 280 (196) | 81.9 (63.1) |
| Quartile 2 | 61.5 (23.4) | 393 (132) | 1267 (144) | 970 (147) | 496 (182) |
| Quartile 3 | 199 (60.6) | 747 (49.7) | 1894 (221) | 1539 (230) | 1110 (176) |
| Quartile 4 | 690 (470) | 1343 (598) | 2907 (488) | 2924 (692) | 2401 (727) |
| **Household characteristics** |  |  |  |  |  |
| Household size, Mean (SD) [Range] | 5.2 (2.6) [2, 18] | 3.8 (1.5) [1, 10] | 4.6 (1.8) [2, 12] | 3.5 (1.5) [1, 10] | 4.3 (2.0) [1, 18] |
| Household wealth at national quintiles, N (%) |  |  |  |  |  |
| 1 (lowest) | 0 | 179 (22%) | 618 (77%) | 114 (14%) | 911 (29%) |
| 2 | 606 (76%) | 401 (50%) | 120 (15%) | 160 (20%) | 1287 (40%) |
| 3 | 155 (19%) | 176 (22%) | 55 (7%) | 189 (24%) | 575 (18%) |
| 4 | 35 (4%) | 43 (5%) | 5 (1%) | 241 (31%) | 324 (10%) |
| 5 (highest) | 0 | 0 | 0 | 89 (11%) | 89 (3%) |
| Missing | 4 (1%) | 0 | 0 | 0 | 4 (<1%) |
| Household primary fuel, N (%) |  |  |  |  |  |
| Charcoal | 0 | 0 | 0 | 193 (24%) | 193 (6%) |
| Cow dung | 0 | 0 | 697 (87%) | 0 | 697 (22%) |
| Wood | 793 (99%) | 799 (100%) | 90 (11%) | 580 (73%) | 2262 (71%) |
| Other | 3 (<1%) | 0 | 10 (1%) | 18 (2%) | 31 (1%) |
| Missing | 4 (<1%) | 0 | 1 (<1%) | 2 (<1%) | 7 (<1%) |
| Household primary cook, N (%) |  |  |  |  |  |
| Pregnant women | 676 (85%) | 757 (95%) | 455 (57%) | 732 (92%) | 2620 (82%) |
| Others in the household | 123 (15%) | 42 (5%) | 342 (43%) | 59 (7%) | 566 (18%) |
| Missing | 1 (<1%) | 0 | 1 (<1%) | 2 (<1%) | 4 (<1%) |
| Someone in the household smokes, N (%) |  |  |  |  |  |
| Yes | 44 (5%) | 253 (32%) | 7 (1%) | 30 (4%) | 334 (10%) |
| No | 756 (95%) | 546 (68%) | 789 (99%) | 761 (96%) | 2852 (89%) |
| Missing | 0 | 0 | 2 (<1%) | 2 (<1%) | 4 (<1%) |

**Note:**
<sup>1</sup> 5 pregnant women on antihypertensive medication are excluded.
<sup>2</sup> MET: Metabolic equivalent of task

### 3.2 Gestational blood pressure

The mean (SD) SBP and DBP in this cohort were 104.8 (9.7) mmHg and 60.7 (7.8) mmHg (**Table 2, Figure S3**). Participants in Rwanda had both the highest mean SBP and DBP; on average, Peruvian participants had the lowest SBP and DBP. Based on the blood pressure classification of American Heart Association, American College of Cardiology (AHA/ACC) Guideline, 93% (2959) of the participants had normal blood pressure (<120/<80 mmHg) and 5% (152) had elevated blood pressure (120-129/<80 mmHg). 56 (2%) were classified as High Blood Pressure (Stage 1) (130-139/80-89 mmHg) and very few (9, <1%) were in the High Blood Pressure (Stage 2) (≥140/≥90 mmHg) category. PP, the difference between SBP and DBP, is a surrogate measure of arterial compliance, with increased PP an indication of arterial stiffness (50,51). On average, participants in Rwanda had the highest PP compared to the other three IRCs. MAP is the average arterial pressure throughout one cardiac cycle and is influenced by cardiac output and systemic vascular resistance (52). 98% of the pregnant women had normal MAP between 60 and 100 mmHg (52,53).

**Table 2.** Measured blood pressure at baseline by IRC

|  | **Guatemala**<br>N = 798 | **India**<br>N = 799 | **Peru**<br>N = 788 | **Rwanda**<br>N = 791 | **All**<br>N = 3176 |
| --- | --- | --- | --- | --- | --- |
| **SBP** |  |  |  |  |  |
| Mean, (SD) | 103.9 (8.5) | 104.5 (9.1) | 99.6 (7.9) | 111.2 (9.4) | 104.8 (9.7) |
| [Range] | [77.7, 145] | [80.0, 142.0] | [74.0, 136.0] | [86.0, 156.0] | [74.0, 156.0] |
| **DBP** |  |  |  |  |  |
| Mean, (SD) | 59.6 (7.2) | 61.5 (7.6) | 57.1 (6.9) | 64.8 (7.4) | 60.7 (7.8) |
| [Range] | [38.5, 88.7] | [42.0, 95.3] | [38.0, 86.0] | [44.0, 112.0] | [38.0, 112.0] |
| **PP** |  |  |  |  |  |
| Mean, (SD) | 44.3 (5.9) | 43.1 (7.4) | 42.5 (6.5) | 46.4 (7.3) | 44.1 (7.0) |
| [Range] | [25.0, 63.0] | [19.3, 67.0] | [21.3, 80.3] | [22.3, 75.7] | [19.3, 80.3] |
| **MAP** |  |  |  |  |  |
| Mean, (SD) | 74.3 (7.1) | 75.8 (7.4) | 71.3 (6.6) | 80.2 (7.4) | 75.4 (7.8) |
| [Range] | [52.8, 105.0] | [56.8, 105.0] | [52.7, 99.9] | [58.0, 125.0] | [52.7, 124.7] |

**Note:**
<sup>1</sup> 14 missing blood pressure measurement at baseline: 2 in Guatemala, 10 in Peru, 2 in Rwanda

### 3.3 Personal exposures to PM_2.5_, BC, and CO

Of the 3190 pregnant women in our analytical population, 2818 (88%), 2536 (79%), and 2872 (90%) participants had valid personal exposure measurements to PM_2.5_, BC, and CO, respectively. Distributions of PM_2.5_, BC, and CO exposure by IRCs are shown in **Figure 1**. The median (IQR) 24-hour PM_2.5_ personal exposure in this cohort is 82.9 (45.9, 145.7) μg/m^3^, and 82% of the participants’ exposures to PM_2.5_ were higher than the World Health Organization’s annual interim target 1 guideline value of 35 μg/m^3^. The median (IQR) of the 24-hour personal exposure to BC and CO was 10.7 (6.5, 15.4) μg/m^3^ and 1.2 (0.5, 2.8) ppm, respectively. Descriptive summaries of exposure to PM_2.5_, BC, and CO after removing the highest and lowest 1% and 5% data points are presented in **Table S1**. Based on the intraclass correlation coefficient for PM_2.5_ (ICC = 0.04), BC (ICC = 0.01), and CO (ICC = 0.05), we observed high within-IRC variability relative to total variability in PM_2.5_/BC/CO exposures. ICCs from models excluding the highest and the lowest 5% of exposures were similar for PM_2.5_ (ICC = 0.05), BC (ICC = 0.01) and CO (ICC = 0.04).

**Figure 1.**
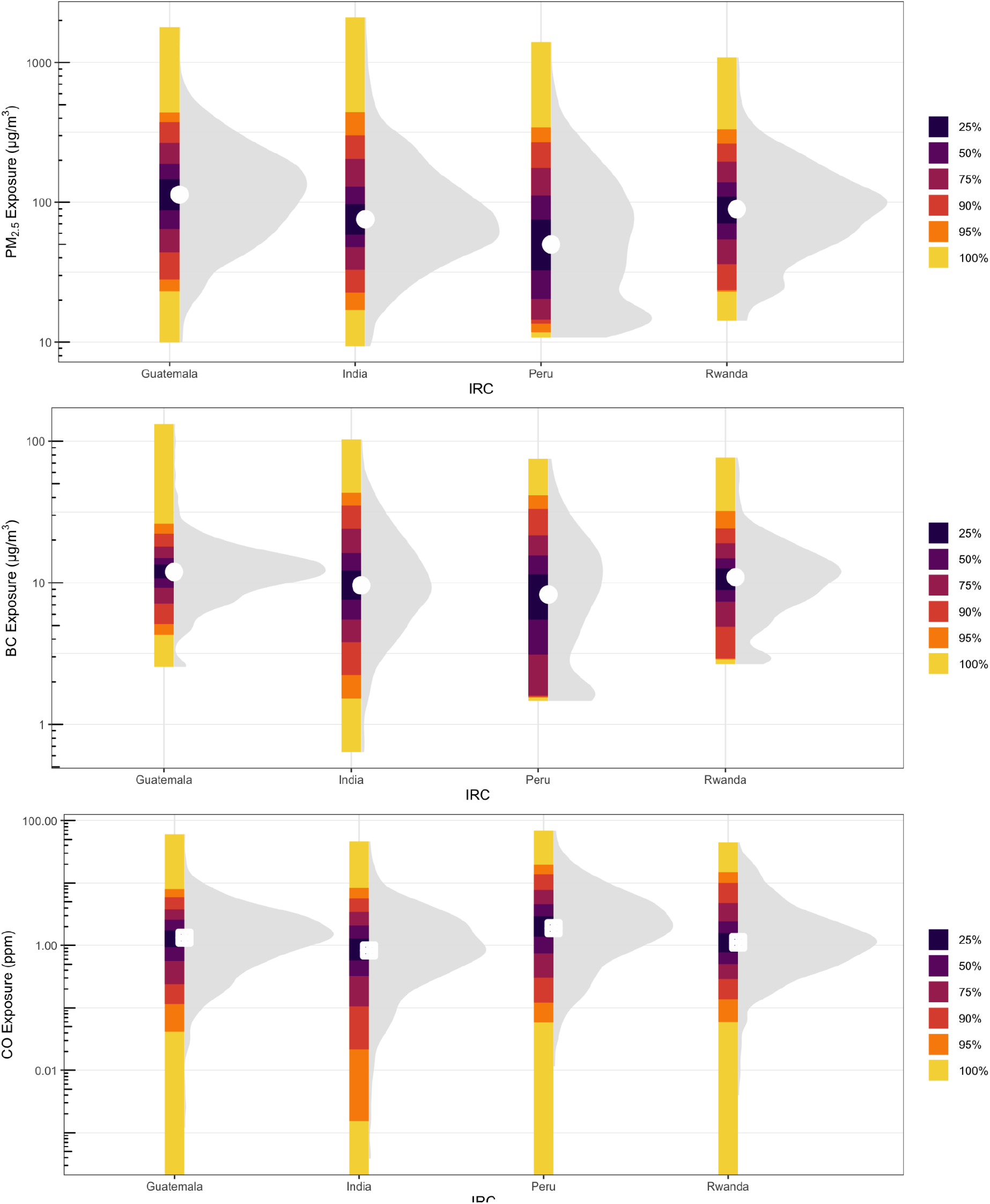
PM_2.5_ (top), BC (middle), and CO (bottom) exposure distributions at baseline by IRC. Shaded areas are density plots. White points are mean values. Vertical bars show the percentage of exposures falling with a given range. Y-axes are log_10_-transformed.

### 3.4 Associations between household air pollution and gestational blood pressure

Analyses were restricted to individuals with both valid exposure and valid blood pressure measurements. Blood pressure and key baseline characteristics (i.e., maternal Age, BMI, nulliparity and mother’s highest education) were generally similar between with and without missing exposure data. Participants with missing PM_2.5_ or CO exposures had relatively lower SBP compared to those with valid exposures. SBP did not differ between missing and non-missing BC exposure participants. Detailed summary statistics are presented in **Table S12**.

The associations between SBP/DBP and covariates (i.e., gestational age, nulliparity, BMI, maternal age, mother’s highest education level, date/time of the blood pressure measurement, and mother’s diet diversity, household food insecurity) were consistent with linearity in each IRC. **Figure S4 – S7** show adjusted associations between PM_2.5_/BC/CO exposures and SBP/DBP in each IRC using GAMs with thin plate regression splines and 3 degrees of freedom. The associations between GBP and HAP are near-linear when considering 95% of exposure samples, as shown in the figures. Judging by AIC, log-linear exposure models generally fit as well as or better than linear exposure models, although for some pollutant/blood pressure associations a categorical exposure model fits better. Therefore, we report our results using log-linear and categorical (quartiles) exposures for all exposure observations (main analysis), and further excluding the highest and the lowest 2.5% of exposures (sensitivity analysis).

Results of trial-wide adjusted associations between PM_2.5_/BC/CO and SBP/DBP using all valid observations are presented in **Table 4**. We did not observe any significant pooled associations between PM_2.5_/BC/CO and SBP/DBP. Positive but non-significant associations are observed in all three pollutants with SBP in log linear exposure models. For DBP, we observed an inverse relationship with higher PM_2.5_/BC/CO exposure associated with lower DBP. However, none of these associations reached conventional statistical significance (*α* = 0.05). We believe the inconsistent direction and magnitude of IRC-specific associations lead to these results (**Figure 2**).

**Table 3.** Measured 24-hour personal exposures to PM_2.5_, BC, and CO (all valid samples) by IRC

| Guatemala | India | Peru | Rwanda | All |
| --- | --- | --- | --- | --- |

| **PM<sub>2.5</sub> exposure (µg/m<sup>3</sup>)** |  |  |  |  |  |
| --- | --- | --- | --- | --- | --- |
| N (% of valid) | 733 (92%) | 715 (89%) | 658 (82%) | 712 (90%) | 2818 (88%) |
| Median | 113.0 | 75.5 | 49.9 | 89.2 | 82.9 |
| (25<sup>th</sup> and 75<sup>th</sup> percentile) | (64.5, 189.0) | (47.7, 129.9) | (20.4, 111.9) | (54.3, 139.1) | (45.9, 145.7) |
| Mean [Range] | 146.0 [9.9, 1799] | 115.0 [9.4, 2100] | 84.9 [10.7, 1400] | 112 [14.2, 1090] | 115.5 [9.4, 2100] |
| **BC exposure (µg/m<sup>3</sup>)** |  |  |  |  |  |
| N (% of valid) | 675 (84%) | 699 (87%) | 596 (75%) | 566 (71%) | 2536 (79%) |
| Median | 11.9 | 9.6 | 8.3 | 10.9 | 10.7 |
| (25<sup>th</sup> and 75<sup>th</sup> percentile) | (9.2, 15.0) | (5.5, 16.2) | (3.1, 15.6) | (7.4, 14.9) | (6.5, 15.4) |
| Mean [Range] | 13.2 [2.6, 133.0] | 12.9 [0.6, 103.0] | 11.3 [1.5, 75.3] | 12.3 [2.7, 76.9] | 12.5 [0.6, 132.6] |
| **CO exposure (ppm)** |  |  |  |  |  |
| N (% of valid) | 757 (95%) | 745 (93%) | 659 (83%) | 711 (90%) | 2872 (90%) |
| Median | 1.3 | 0.8 | 1.9 | 1.1 | 1.2 |
| (25<sup>th</sup> and 75<sup>th</sup> percentile) | (0.6, 2.6) | (0.3, 2.1) | (0.9, 4.6) | (0.5, 2.4) | (0.5, 2.8) |
| Mean [Range] | 2.0 [0, 60.2] | 1.8 [0, 46.9] | 3.9 [0, 69.5] | 2.5 [0, 44.4] | 2.5 [0, 69.5] |

**Table 4.** Trial-wide adjusted association between HAP exposure and gestational blood pressure (all valid samples).

| | $\text{PM}_{2.5}$ | | BC | | CO | |
| --- | --- | --- | --- | --- | --- | --- |
|  | Estimate | 95% CI | Estimate | 95% CI | Estimate | 95% CI |
| ***Systolic Blood Pressure*** |  |  |  |  |  |  |
| Log linear | 0.04 | (-0.35, 0.43) | 0.15 | (-0.31, 0.62) | 0.01 | (-0.30, 0.33) |
| Categorical [Ref. Quartile 1] |  |  |  |  |  |  |
| Quartile 2 | -0.07 | (-0.99, 0.86) | -0.67 | (-1.60, 0.26) | -0.44 | (-2.31, 1.42) |
| Quartile 3 | 0.08 | (-0.85, 1.02) | -0.41 | (-1.98, 1.15) | 0.66 | (-0.44, 1.76) |
| Quartile 4 | 0.26 | (-0.68, 1.20) | 0.29 | (-0.85, 1.43) | -0.32 | (-1.47, 0.82) |
| ***Diastolic Blood Pressure*** |  |  |  |  |  |  |
| Log linear | -0.29 | (-0.95, 0.38) | -0.24 | (-0.91, 0.43) | -0.01 | (-0.36, 0.33) |
| Categorical [Ref. Quartile 1] |  |  |  |  |  |  |
| Quartile 2 | -0.12 | (-0.88, 0.64) | -0.35 | (-1.15, 0.44) | -0.66 | (-2.16, 0.84) |
| Quartile 3 | -0.41 | (-1.84, 1.01) | -0.60 | (-2.10, 0.91) | -0.07 | (-1.26, 1.11) |
| Quartile 4 | -0.22 | (-1.53, 1.10) | -0.70 | (-2.19, 0.78) | -0.08 | (-1.70, 1.54) |

**Notes:**
1. All models controlled for gestational age at the BP measurement, BMI, and mother's age. Additional covariates are controlled for in IRC-specific models (Guatemala: nulliparity, mother's education, physical activity, date of the BP measurement, mother's diet diversity and season; India: mother's education, household wealth, and season; Peru: physical activity, time of the BP measurement, household food insecurity, mother's diet diversity and season; Rwanda: nulliparity, mother's education, physical activity, time of the BP measurement, household food insecurity and smoker at home) 2. In log-linear models, the coefficients indicate the increase in BP (mmHg) per a one unit increase in log exposure. 3. Shaded cells are fixed effects, unshaded are random effects, meta-analyses combining results across 4 IRCs

**Figure 2.**
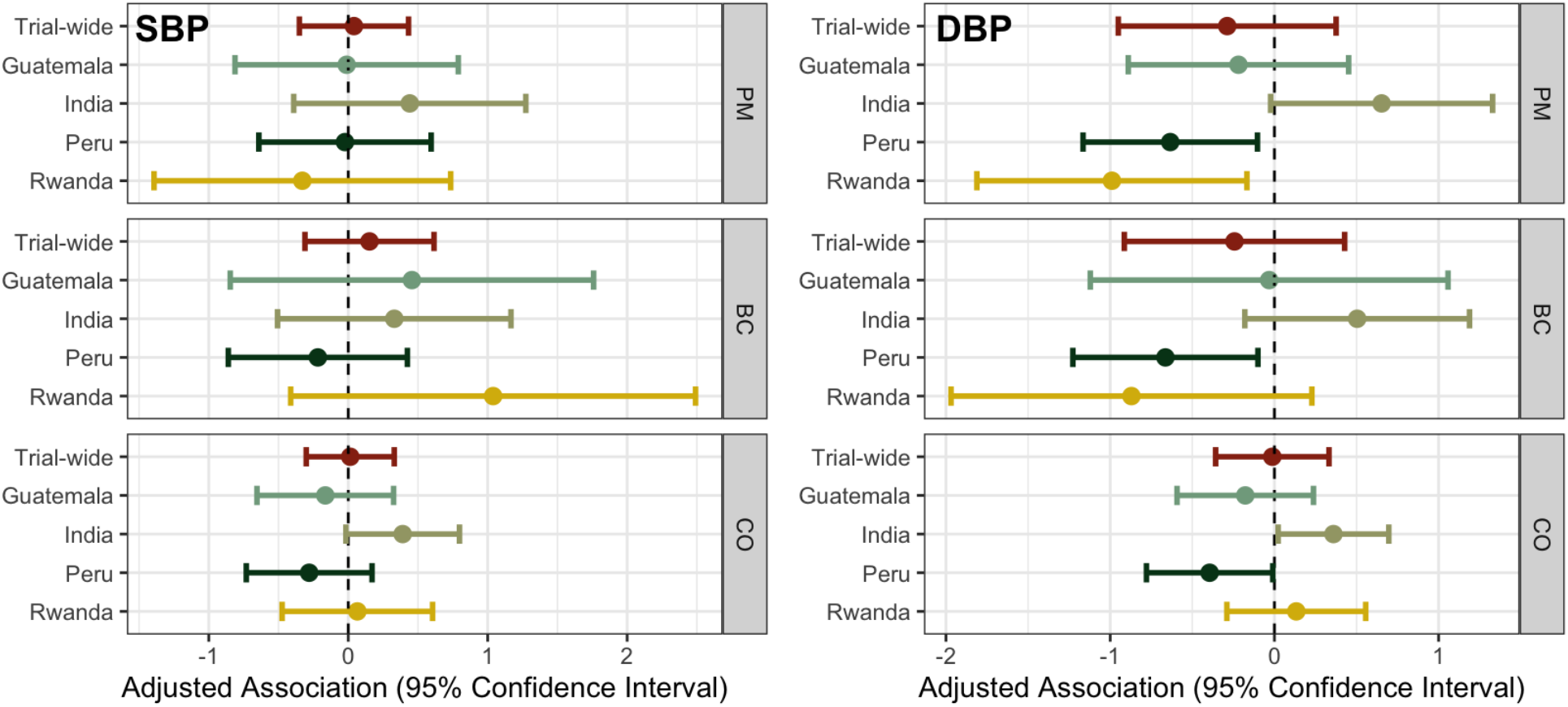
Trial-wide and IRC-specific adjusted associations and 95% confidence interval between PM_2.5_/BC/CO exposure and SBP/DBP in log-linear exposure models. Colors are used to distinguish between trial-wide results and IRC-specific results.

Distinct from the other three IRCs, India showed an consistent trends with increased SBP/DBP for higher exposures across all three measured pollutants, as reported previously (42). Briefly, we observed a positive association between CO and DBP among the participants in India: a 1-log µg/m^3^ increase in CO exposure was associated with 0.36 (95% CI: 0.02, 0.70) mmHg higher DBP. In Guatemala, except for BC and SBP, all the other associations are negative, and none of these associations reached statistical significance. In Peru, all HAP exposure and blood pressure associations were negative. Specifically, there were significant associations between HAP exposures and DBP: a 1-log µg/m^3^ increase in PM_2.5_, BC and CO was associated with −0.63 (95% CI: −1.17, −0.10) mmHg, −0.66 (95% CI: −1.23, −0.10) mmHg, and −0.39 (95% CI: −0.78, −0.01) mmHg change in DBP, respectively. In Rwanda, non-significant increases in SBP and DBP were related to increased exposure to CO in log-linear exposure models. The associations between PM_2.5_/BC and SBP/DBP were inconsistent in direction and a significant inverse association between PM_2.5_ and DBP was found: 1-log µg/m^3^ increase in PM_2.5_ was associated with −0.99 (95% CI: −1.81, −0.17) mmHg lower DBP.

The relationships between PM_2.5_/BC/CO exposures and PP/MAP showed similar pattern. Detailed trial-wide and IRC-specific associations of these secondary endpoints are presented in **Table S2** and **Table S3b – S6b**. There was some evidence to suggest an increase in PP with higher PM_2.5_ and BC exposures in Peru and Rwanda.

Removing the 5% of the extreme exposure observations resulted in similar trends and magnitudes of the associations between PM_2.5_/BC/CO exposure and all blood pressure parameters (**Table S7**). Gestational age at blood pressure measurement, physical activity, and the presence of smoker at home did not modify the associations between PM_2.5_/BC/CO exposure and SBP/DBP across all IRCs. We observed evidence that BMI modified the associations with SBP in India and Peru, with stronger associations among obese women (BMI ≥ 30 kg/m^2^) compared to women who were considered normal or healthy weight (BMI between 18.5 and 24.9 kg/m^2^). Among obese women, a 1-log µg/m^3^ increase in PM_2.5_ exposure was associated with 7.39 (95% CI: 1.10, 13.68) mmHg higher SBP in India and 2.36 (95% CI: 0.42, 4.29) mmHg higher SBP in Peru; a 1-log µg/m^3^ increase in BC was associated with 14.23 (95% CI: 4.12, 24.34) mmHg higher SBP in India and 2.26 (95% CI: 0.20, 4.32) mmHg higher SBP in Peru. We did not see similar trends in DBP. Detailed effect modification results can be found in **Table S8 – S11**.

## 4. DISCUSSION

As part of the HAPIN RCT baseline measurement period, we conducted PM_2.5_, BC, and CO personal exposure assessment (n = 2818, 2536, 2872, respectively) and collected gestational blood pressure during early pregnancy (n = 3176, average of 15.4 gestational weeks). Exposure levels were consistently above recommended WHO Interim Target values; blood pressure values varied by IRC but were generally within normotensive ranges (n = 2959, 93%).

Trial-wide, among pregnant women, we observed higher SBP but lower DBP with higher exposures to PM_2.5_/BC/CO; however, none of the associations reached conventional statistical significance at *α* = 0.05. This result is driven by the inconsistent associations (in both direction and magnitude) within each IRC. Specifically, we observed negative associations between all exposures and SBP/DBP in Guatemala, except for BC and SBP relationship; none of the associations was significant. In India, higher blood pressure was associated with higher HAP exposure; a significant association was found between CO and DBP: a 1-log µg/m^3^ increase in CO exposure was associated with 0.36 (95% CI: 0.02, 0.70) mmHg higher DBP. On the contrary, in Peru, lower blood pressure was seen in higher exposure participants; in this case, the relationship between PM_2.5_/BC/CO exposure and DBP reached conventional statistical significance. None of the HAP exposure and blood pressure parameters were significantly associated in Rwanda and the directions were inconsistent.

Associations between HAP exposure and blood pressure reported in the literature vary. Among older, non-pregnant women (15,16,28), evidence of an association exists between HAP exposure and SBP, DBP, or both. The comparison between older adult women and pregnant women may, however, not be appropriate due to both physiological and risk factor-related differences. Two studies (29,30) evaluated the impacts of HAP on gestational blood pressure and found positive associations between HAP exposure and DBP, though only one presented quantitative exposure-response evidence for CO (30).

Our findings, though inconsistent with the existing literature base for pregnant women, were robust across model specifications and sensitivity analyses. Using a common personal exposure and blood pressure assessment protocol across four diverse biomass-using LMIC populations, this analysis also demonstrated the heterogeneity in the associations between HAP exposure and blood pressure among pregnant women in early-to mid-pregnancy. The unexpected cross-sectional inverse association between PM_2.5_/BC/CO exposures and DBP in Guatemala, Peru and Rwanda might be caused by unmeasured preexisting and pregnancy-related factors that had greater impact on gestational blood pressure than HAP exposures, such as diet and nutrition, family health history and gestational weight gain (35,54,55). However, most of these associations were not statistically significant. For those that were, the size of the associations was all < 1 mmHg and would likely not be considered clinically significant (56).

Furthermore, we did not observe similar decreases in SBP in the contexts where we noted decreases in DBP, leading to increases in PP with higher exposures. Increased PP is an indicator of reduced arterial compliance and has been proposed as a predictor of hypertensive disorders of pregnancy when elevated in early pregnancy (46,50). However, the association between HAP exposure and PP was only significant in Peru: 1-log µg/m^3^ increase in PM_2.5_ was associated with 0.61 (95% CI: 0.09, 1.13) mmHg increase in PP.

Additionally, we observed evidence that BMI modified the associations between PM_2.5_/BC and SBP in India and Peru. A 7.39 (95% CI: 1.10, 13.68) and 2.36 (95% CI: 0.42, 4.29) mmHg SBP increase was observed among obese women compared to normal or healthy weight women for 1-log µg/m^3^ increase in PM_2.5_ in India and Peru, respectively. We also saw a statistically significant interaction between BC exposure and BMI for SBP. Compared to women who were considered normal or healthy weight, we found a 14.23 (95% CI: 4.12, 24.34) and 2.26 (95% CI: 0.20, 4.32) mmHg higher SBP among obese for 1 log µg/m3 increase in BC exposure. However, given the small number of obese women in India, the observed interaction effect of BMI should be interpreted with caution.

Given the paucity of data on the impact of HAP on gestational blood pressure – and the inconsistency in findings – further evaluation is likely needed. Recent evidence from China (57) indicates that the magnitude and trajectory of changes in blood pressure during pregnancy vary by quartile of exposure to ambient PM_2.5_. That is, the trajectory of typical changes in blood pressure during pregnancy are altered by ambient PM exposure. Explorations of such changes in trajectory from HAP exposure may be valuable and would benefit from a repeat measurement strategy, which captures both BP and exposure throughout pregnancy, as undertaken during the broader HAPIN trial.

Our study has several strengths. We performed high quality personal exposure and blood pressure measurement among pregnant women in four diverse low- and middle-income settings. As planned for the trial, baseline data was collected relatively early during pregnancy. We also acknowledge a number of limitations. First, this cross-sectional analysis assessed HAP exposure and blood pressure at a single time point in early pregnancy. Findings from additional rounds of measurement during the HAPIN trial are being prepared. Second, single measurements of both blood pressure and HAP exposure are known to be variable, and thus some amount of measurement error is expected. Third, this analysis focuses only on the HAPIN baseline period, when all households were cooking with biomass; we do not benefit from additional potential heterogeneity in exposure due to the HAPIN stove, fuel, and behavioral intervention.

Forthcoming evaluation of the effect of the HAPIN intervention on blood pressure among both pregnant and older adult women will help elucidate the relationship between household air pollution exposure and blood pressure. Given the burden of ill-health associated with elevated blood pressure and related outcomes, further investigation of its relationship with HAP exposure is warranted.

## 5. CONCLUSIONS

In summary, this study added new evidence to the understanding of the association between HAP exposure and blood pressure in pregnant women. Our analysis of 3190 young and low risk pregnant women from four diverse LMICs demonstrated the heterogeneity in the associations between HAP exposure and blood pressure during early-to mid-pregnancy and provided evidence of an effect modification by BMI. These findings motivate future longitudinal studies among populations with different risk factors of pregnancy.

## Data Availability

All data produced in the present study are available upon reasonable request to the authors

## Acknowledgements

The HAPIN trial is funded by the U.S. National Institutes of Health (cooperative agreement 1UM1HL134590) in collaboration with the Bill & Melinda Gates Foundation (OPP1131279). The investigators would like to thank the members of the advisory committee - Patrick Brysse, Donna Spiegelman, and Joel Kaufman - for their valuable insight and guidance throughout the implementation of the trial. We also wish to acknowledge all research staff and study participants for their dedication to and participation in this important trial.

A multidisciplinary, independent Data and Safety Monitoring Board (DSMB) appointed by the National Heart, Lung, and Blood Institute (NHLBI) monitors the quality of the data and protects the safety of patients enrolled in the HAPIN trial. NHLBI DSMB: Nancy R Cook, Stephen Hecht, Catherine Karr (Chair), Joseph Millum, Nalini Sathiakumar, Paul K Whelton, Gail Weinmann and Thomas Croxton (Executive Secretaries). Program Coordination: Gail Rodgers, Bill & Melinda Gates Foundation; Claudia L Thompson, National Institute of Environmental Health Science; Mark J. Parascandola, National Cancer Institute; Marion Koso-Thomas, Eunice Kennedy Shriver National Institute of Child Health and Human Development; Joshua P Rosenthal, Fogarty International Center; Conception R Nierras, NIH Office of Strategic Coordination Common Fund; Katherine Kavounis, Dong-Yun Kim, Antonello Punturieri, and Barry S Schmetter, NHLBI. The findings and conclusions in this report are those of the authors and do not necessarily represent the official position of the US National Institutes of Health or Department of Health and Human Services.

## Data Sharing

|  |  |
| --- | --- |
| Will individual participant data be available (including data dictionaries)? | Yes |
| What data in particular will be shared? | Individual participant data that underlie the results reported in this article, after de-identification (all results summarized in text, tables, figures, and appendices) |
| What other documents will be available? | Study protocol, statistical analysis plan, informed consent form, analytic code |
| When will data be available (start and end dates)? | Beginning 6 months following article publication |
| With whom? | Anyone who wishes to access the data |
| For what types of analyses? | Any purpose |
| By what mechanism will data be made available? | Data are available indefinitely at DataVerse (link to be included) |

## Authors’ contribution

Conceptualization, Wenlu Ye and Ajay Pillarisetti; Data curation, Yunyun Chen and Jiantong Wang; Formal analysis, Wenlu Ye, Ajay Pillarisetti and Oscar De León; Investigation, Kyle Steenland and Jennifer Peel; Methodology, Wenlu Ye, Ajay Pillarisetti, Kyle Steenland and Jennifer Peel; Resources, Lindsay Underhill, Kalpana Balakrishnan, John McCracken, Lisa Thompson, Anaite Diaz-Artiga, Ghislaine Rosa, Victor Davila-Roman and Lisa De las Fuentes; Software, Wenlu Ye, Ajay Pillarisetti and Oscar De León; Supervision, Ajay Pillarisetti; Visualization, Wenlu Ye and Ajay Pillarisetti; Writing – original draft, Wenlu Ye and Ajay Pillarisetti; Writing – review & editing, Kyle Steenland, Jennifer Peel, Maggie Clark, William Checkley, Ashlinn Quinn, Kalpana Balakrishnan, Ghislaine Rosa, Aris Papageorghiou, Sarada Garg and Thomas Clasen.

## Declaration of interests

The authors declare that they have no known competing financial interests or personal relationships that could have appeared to influence the work reported in this paper.

## HAPIN Investigators

Vigneswari Aravindalochanan, Kalpana Balakrishnan, Gloriose Bankundiye, Dana Boyd Barr, Vanessa Burrowes, Alejandra Bussalleu, Devan Campbell, Eduardo Canuz, Adly Castañaza, Howard H. Chang, William Checkley, Yunyun Chen, Marilú Chiang, Maggie L. Clark, Thomas F. Clasen, Rachel Craik, Mary Crocker, Victor G. Davila-Roman, Lisa de las Fuentes, Oscar De León, Anaité Diaz-Artiga, Ephrem Dusabimana, Lisa Elon, Juan Gabriel Espinoza, Irma Sayury Pineda Fuentes, Sarada Garg, Ahana Ghosh, Dina Goodman, Savannah Gupton, Sarah Hamid, Stella Hartinger, Steven A. Harvey, Mayari Hengstermann, Ian Hennessee, Phabiola Herrera, Shakir Hossen, Marjorie Howard, Penelope P. Howards, Lindsay Jaacks, Shirin Jabbarzadeh, Michael A. Johnson, Katherine Kearns, Miles A. Kirby, Jacob Kremer, Margaret A. Laws, Pattie Lenzen, Jiawen Liao, Amy E. Lovvorn, Jane Mbabazi, Eric McCollum, John P. McCracken, Julia N. McPeek, Rachel Meyers, J. Jaime Miranda, Erick Mollinedo, Libny Monroy, Lawrence Moulton, Alexie Mukeshimana, Krishnendu Mukhopadhyay, Bernard Mutariyani, Luke Naeher, Abidan Nambajimana, Durairaj Natesan, Florien Ndagijimana, Laura Nicolaou, Azhar Nizam, Jean de Dieu Ntivuguruzwa, Aris Papageorghiou, Jennifer Peel, Ricardo Piedrahita, Ajay Pillarisetti, Naveen Puttaswamy, Elisa Puzzolo, Ashlinn Quinn, Karthikeyan Dharmapuri Rajamani, Sarah Rajkumar, Usha Ramakrishnan, Rengaraj Ramasami, Alexander Ramirez, Ghislaine Rosa, Joshua Rosenthal, P. Barry Ryan, Sudhakar Saidam, Zoe Sakas, Sankar Sambandam, Jeremy A. Sarnat, Suzanne Simkovich, Sheela S. Sinharoy, Kirk R. Smith, Kyle Steenland, Damien Swearing, Gurusamy Thangavel, Lisa M. Thompson, Ashley Toenjes, Lindsay Underhill, Jean Damascene Uwizeyimana, Viviane Valdes, Amit Verma, Lance A. Waller, Jiantong Wang, Megan Warnock, Kendra N. Williams, Wenlu Ye, Bonnie N. Young, Ashley Younger.

## Notes

### Competing Interest Statement

The authors have declared no competing interest.

### Clinical Trial

NCT02944682

### Clinical Protocols

https://clinicaltrials.gov/ct2/show/NCT02944682

### Author Declarations

Ethics committee/IRB of Emory University gave ethical approval for this work (00089799) Ethics committee/IRB of Johns Hopkins University gave ethical approval for this work (00007403) Ethics committee/IRB of Sri Ramachandra Institute of Higher Education and Research gave ethical approval for this work (IEC–N1/16/JUL/54/49) Ethics committee/IRB of Indian Council of Medical Research – Health Ministry Screening Committee gave ethical approval for this work (5/8/4–30/(Env)/Indo–US/2016–NCD–I) Ethics committee/IRB of Universidad del Valle de Guatemala gave ethical approval for this work (146–08–2016) Ethics committee/IRB of Guatemalan Ministry of Health National Ethics Committee gave ethical approval for this work (11–2016) Ethics committee/IRB of Asociacion Beneficia PRISMA gave ethical approval for this work (CE2981.17) Ethics committee/IRB of London School of Hygiene and Tropical Medicine gave ethical approval for this work (11664–3) Ethics committee/IRB of Rwandan National Ethics Committee gave ethical approval for this work (No.016/RNEC/2018) Ethics committee/IRB of Washington University in St. Louis gave ethical approval for this work (201611159)

