## Supplementary Material for "Baseline associations between household air pollution exposure and blood pressure among pregnant women in the Household Air Pollution Intervention Network (HAPIN) multi-country randomized controlled trial"

**Table of Contents**


| | |
|---|---|
| **Table S1**. Measured 24-hour personal exposures to $PM_{2.5}$, BC, and CO by IRC, after removing the lowest and the highest 2.5% of exposure samples | 2 |
| **Table S2.** Trial-wide adjusted association between HAP exposure and PP/MAP | 2 |
| **Table S3a.** Adjusted association between HAP and SBP/DBP in Guatemala | 3 |
| **Table S3b.** Adjusted association between HAP and PP/MAP in Guatemala | 4 |
| **Table S4a.** Adjusted association between HAP and SBP/DBP in India | 5 |
| **Table S4b.** Adjusted association between HAP and PP/MAP in India | 6 |
| **Table S5a.** Adjusted association between HAP and SBP/DBP in Peru | 7 |
| **Table S5b.** Adjusted association between HAP and PP/MAP in Peru | 8 |
| **Table S6a.** Adjusted association between HAP and SBP/DBP in Rwanda | 9 |
| **Table S6b.** Adjusted association between HAP and PP/MAP in Rwanda | 10 |
| **Table S7.** Trial-wide association between HAP exposure and blood pressure parameters (without the highest and lowest 2.5% of the exposure samples) | 11 |
| **Table S8.** Effect modification for the HAP-BP association in Guatemala | 11 |
| **Table S9.** Effect modification for the HAP-BP association in India | 13 |
| **Table S10.** Effect modification for the HAP-BP association in Peru | 14 |
| **Table S11.** Effect modification for the HAP-BP association in Rwanda | 15 |
| **Table S12**. Summary of SBP, DBP and selected baseline characteristics in population with and without missing exposures to $PM_{2.5}$, BC, and CO | 16 |
| **Figure S1**. Directed Acyclic Graph (DAG) for the HAP-BP association | 17 |
| **Figure S2.** Distributions of gestational age at baseline | 18 |
| **Figure S3.** Distributions of systolic (top) and diastolic (bottom) blood pressure at baseline | 19 |
| **Figure S4.** HAP-GBP association and 95% confidence intervals in Guatemala | 20 |
| **Figure S5.** HAP-GBP association and 95% confidence intervals in India | 21 |
| **Figure S6.** HAP-GBP association and 95% confidence intervals in Peru | 22 |
| **Figure S7.** HAP-GBP association and 95% confidence intervals in Rwanda | 23 |

**Table S1**. Measured 24-hour personal exposures to $PM_{2.5}$, BC, and CO by IRC, after removing the lowest and the highest 2.5% of exposure samples

| | **N** | **Median** | **IQR** | **Mean** | **Min** | **Max** |
|---|---|---|---|---|---|---|
| ***PM2.5 > 14.25 and PM2.5 < 401.55 (removed highest and lowest 2.5%)*** | | | | | | |
| All IRCs | 2676 | 82.85 | 93.75 | 104.80 | 14.25 | 400.05 |
| Guatemala | 699 | 110.40 | 114.00 | 130.40 | 15.13 | 392.30 |
| India | 685 | 75.08 | 78.85 | 97.94 | 15.85 | 388.60 |
| Peru | 591 | 54.94 | 89.24 | 82.63 | 14.25 | 378.50 |
| Rwanda | 701 | 88.35 | 81.26 | 104.70 | 14.29 | 400.10 |
| ***BC > 1.57 and BC < 39.30 (removed highest and lowest 2.5%)*** | | | | | | |
| All IRCs | 2408 | 10.69 | 8.20 | 11.61 | 1.57 | 38.83 |
| Guatemala | 665 | 11.90 | 5.70 | 12.37 | 2.55 | 34.30 |
| India | 656 | 9.41 | 9.94 | 11.72 | 1.58 | 38.39 |
| Peru | 531 | 8.72 | 11.37 | 10.69 | 1.57 | 38.83 |
| Rwanda | 556 | 10.87 | 7.42 | 11.46 | 2.66 | 35.97 |
| ***CO > 0.03 and CO < 13.63 (removed highest and lowest 2.5%)*** | | | | | | |
| All IRCs | 2728 | 1.24 | 2.13 | 2.03 | 0.03 | 13.63 |
| Guatemala | 741 | 1.36 | 2.09 | 1.94 | 0.03 | 12.61 |
| India | 697 | 0.88 | 1.73 | 1.58 | 0.03 | 12.15 |
| Peru | 616 | 1.81 | 3.33 | 2.84 | 0.03 | 13.63 |
| Rwanda | 674 | 1.08 | 1.71 | 1.86 | 0.03 | 13.56 |

**Table S2.** Trial-wide adjusted association between HAP exposure and PP/MAP.

| | $PM_{2.5}$ | | BC | | CO | |
|---|---|---|---|---|---|---|
| | Estimate | 95% CI | Estimate | 95% CI | Estimate | 95% CI |
| ***Pulse Pressure (PP)*** | | | | | | |
| Log linear | 0.32 | (-0.05, 0.70) | 0.58 | (-0.14, 1.29) | 0.03 | (-0.15, 0.21) |
| Categorical [Ref. Quartile 1] | | | | | | |
| Quartile 2 | 0.07 | (-0.64, 0.78) | -0.34 | (-1.09, 0.41) | 0.13 | (-0.56, 0.83) |
| Quartile 3 | 0.49 | (-0.42, 1.41) | 0.21 | (-0.62, 1.05) | 0.73 | (0.03, 1.42) |
| Quartile 4 | 0.52 | (-0.20, 1.25) | 1.04 | (-0.06, 2.14) | -0.19 | (-0.89, 0.52) |
| ***Mean Arterial Pressure (MAP)*** | | | | | | |
| Log linear | -0.18 | (-0.71, 0.36) | -0.07 | (-0.59, 0.45) | -0.01 | (-0.34, 0.33) |
| Categorical [Ref. Quartile 1] | | | | | | |
| Quartile 2 | -0.12 | (-0.86, 0.63) | -0.45 | (-1.23, 0.33) | -0.58 | (-2.20, 1.03) |
| Quartile 3 | -0.25 | (-1.37, 0.87) | -0.52 | (-1.94, 0.91) | 0.18 | (-0.96, 1.32) |
| Quartile 4 | -0.05 | (-1.14, 1.03) | -0.35 | (-1.63, 0.93) | -0.15 | (-1.62, 1.32) |

***Note:***
*1. All models controlled for gestational age at the BP measurement, BMI, and mother's age. Additional covariates are controlled for IRC-specific models (Guatemala: nulliparity, mother's education, physical activity, date of the BP measurement, mother's diet diversity and season; India: mother's education, household wealth, and season; Peru: physical activity, time of the BP measurement, household food insecurity, mother's diet diversity and season; Rwanda: nulliparity, mother's education, physical activity, time of the BP measurement, household food insecurity and smoker at home)*
*2. In log linear models, the coefficients indicate the increase in BP (mmHg) per a one unit increase in the log of exposure.*

*3. Shaded cells are fixed effects, unshaded are random effects, meta-analyses combining results across 4 IRCs*

**Table S3a.** Adjusted association between HAP and SBP/DBP in Guatemala

| | Model Type | Estimate | p-value | 95% CI | AIC |
|---|---|---|---|---|---|
| *Systolic Blood Pressure* | | | | | |
| PM2.5 | Linear | 3.00E-04 | 0.9069 | (-0.0046, 0.0051) | 5139 |
| | Log linear | -0.0115 | 0.9776 | (-0.8122, 0.7892) | 5139 |
| | Categorical [Ref. Quartile 1) | | | | |
| | Quartile 2 | 0.7817 | 0.3727 | (-0.9388, 2.5022) | 5142 |
| | Quartile 3 | 0.4535 | 0.6087 | (-1.2849, 2.192) | 5142 |
| | Quartile 4 | 0.5567 | 0.5318 | (-1.1906, 2.3039) | 5142 |
| BC | Linear | 0.028 | 0.4141 | (-0.0392, 0.0952) | 4700 |
| | Log linear | 0.4565 | 0.4917 | (-0.8464, 1.7593) | 4700 |
| | Categorical [Ref. Quartile 1) | | | | |
| | Quartile 2 | -0.2543 | 0.7779 | (-2.024, 1.5154) | 4704 |
| | Quartile 3 | -0.1518 | 0.8662 | (-1.9199, 1.6164) | 4704 |
| | Quartile 4 | 0.5654 | 0.5313 | (-1.207, 2.3379) | 4704 |
| CO | Linear | -0.0378 | 0.702 | (-0.2316, 0.156) | 5245 |
| | Log linear | -0.1647 | 0.5092 | (-0.6544, 0.325) | 5245 |
| | Categorical [Ref. Quartile 1) | | | | |
| | Quartile 2 | -2.0871 | 0.0153 | (-3.7726, -0.4015) | 5242 |
| | Quartile 3 | -0.454 | 0.5999 | (-2.1526, 1.2446) | 5242 |
| | Quartile 4 | -0.9296 | 0.2891 | (-2.6501, 0.7909) | 5242 |
| *Diastolic Blood Pressure* | | | | | |
| PM2.5 | Linear | -0.001 | 0.6146 | (-0.0051, 0.003) | 4882 |
| | Log linear | -0.2189 | 0.5221 | (-0.8899, 0.4521) | 4882 |
| | Categorical [Ref. Quartile 1) | | | | |
| | Quartile 2 | 0.6895 | 0.348 | (-0.752, 2.131) | 4885 |
| | Quartile 3 | 0.2093 | 0.778 | (-1.2473, 1.6658) | 4885 |
| | Quartile 4 | -0.1483 | 0.8424 | (-1.6123, 1.3156) | 4885 |
| BC | Linear | 0.0169 | 0.555 | (-0.0393, 0.0731) | 4461 |
| | Log linear | -0.0319 | 0.9542 | (-1.121, 1.0573) | 4461 |
| | Categorical [Ref. Quartile 1) | | | | |
| | Quartile 2 | -0.0011 | 0.9989 | (-1.4807, 1.4785) | 4465 |
| | Quartile 3 | -0.112 | 0.8818 | (-1.5902, 1.3663) | 4465 |
| | Quartile 4 | -0.417 | 0.5808 | (-1.8989, 1.0649) | 4465 |
| CO | Linear | -0.062 | 0.4593 | (-0.2264, 0.1024) | 5001 |
| | Log linear | -0.1775 | 0.4018 | (-0.5929, 0.2379) | 5001 |
| | Categorical [Ref. Quartile 1) | | | | |
| | Quartile 2 | -1.5119 | 0.0386 | (-2.9443, -0.0796) | 5001 |
| | Quartile 3 | -0.7466 | 0.3103 | (-2.19, 0.6969) | 5001 |
| | Quartile 4 | -0.8224 | 0.2698 | (-2.2845, 0.6397) | 5001 |

***Note:*** *All models controlled for gestational age at the BP measurement, BMI, and mother's age, nulliparity, mother's education, physical activity, date of the BP measurement, mother's diet diversity and season*

**Table S3b.** Adjusted association between HAP and PP/MAP in Guatemala

| | Model Type | Estimate | p-value | 95% CI | AIC |
|---|---|---|---|---|---|
| *Pulse Pressure* | | | | | |
| PM2.5 | Linear | 0.0013 | 0.4410 | (-0.0021, 0.0047) | 4620 |
| | Log linear | 0.2074 | 0.4673 | (-0.3526, 0.7674) | 4620 |
| | Categorical [Ref. Quartile 1) | | | | |
| | Quartile 2 | 0.0922 | 0.8804 | (-1.111, 1.2954) | 4623 |
| | Quartile 3 | 0.2443 | 0.6934 | (-0.9715, 1.46) | 4623 |
| | Quartile 4 | 0.7050 | 0.2577 | (-0.5169, 1.9269) | 4623 |
| BC | Linear | 0.0111 | 0.6536 | (-0.0373, 0.0595) | 4263 |
| | Log linear | 0.4883 | 0.3071 | (-0.4498, 1.4264) | 4262 |
| | Categorical [Ref. Quartile 1) | | | | |
| | Quartile 2 | -0.2532 | 0.6959 | (-1.5248, 1.0184) | 4263 |
| | Quartile 3 | -0.0398 | 0.9510 | (-1.3102, 1.2306) | 4263 |
| | Quartile 4 | 0.9824 | 0.1303 | (-0.2911, 2.2559) | 4263 |
| CO | Linear | 0.0242 | 0.7260 | (-0.1114, 0.1598) | 4715 |
| | Log linear | 0.0128 | 0.9416 | (-0.3299, 0.3555) | 4715 |
| | Categorical [Ref. Quartile 1) | | | | |
| | Quartile 2 | -0.5752 | 0.3401 | (-1.758, 0.6077) | 4717 |
| | Quartile 3 | 0.2926 | 0.6301 | (-0.8994, 1.4846) | 4717 |
| | Quartile 4 | -0.1072 | 0.8616 | (-1.3146, 1.1001) | 4717 |
| *Mean Arterial Pressure* | | | | | |
| PM2.5 | Linear | -0.0006 | 0.7709 | (-0.0046, 0.0034) | 4873 |
| | Log linear | -0.1497 | 0.6593 | (-0.8162, 0.5167) | 4873 |
| | Categorical [Ref. Quartile 1) | | | | |
| | Quartile 2 | 0.7203 | 0.3237 | (-0.7116, 2.1521) | 4876 |
| | Quartile 3 | 0.2907 | 0.6934 | (-1.1561, 1.7375) | 4876 |
| | Quartile 4 | 0.0867 | 0.9069 | (-1.3675, 1.5408) | 4876 |
| BC | Linear | 0.0206 | 0.4672 | (-0.035, 0.0761) | 4447 |
| | Log linear | 0.1309 | 0.8115 | (-0.9466, 1.2085) | 4447 |
| | Categorical [Ref. Quartile 1) | | | | |
| | Quartile 2 | -0.0855 | 0.9088 | (-1.5498, 1.3788) | 4451 |
| | Quartile 3 | -0.1252 | 0.8666 | (-1.5882, 1.3378) | 4451 |
| | Quartile 4 | -0.0895 | 0.9046 | (-1.5561, 1.377) | 4451 |
| CO | Linear | -0.0539 | 0.5152 | (-0.2166, 0.1087) | 4985 |
| | Log linear | -0.1733 | 0.4081 | (-0.5842, 0.2377) | 4985 |
| | Categorical [Ref. Quartile 1) | | | | |
| | Quartile 2 | -1.7036 | 0.0184 | (-3.1192, -0.2881) | 4983 |
| | Quartile 3 | -0.6490 | 0.3720 | (-2.0756, 0.7775) | 4983 |
| | Quartile 4 | -0.8581 | 0.2440 | (-2.3031, 0.5868) | 4983 |

***Note:*** *All models controlled for gestational age at the BP measurement, BMI, and mother's age, nulliparity, mother's education, physical activity, date of the BP measurement, mother's diet diversity and season.*

**Table S4a.** Adjusted association between HAP and SBP/DBP in India

| | Model Type | Estimate | p-value | 95% CI | AIC |
|---|---|---|---|---|---|
| *Systolic Blood Pressure* | | | | | |
| PM2.5 | Linear | 0.0023 | 0.3281 | (-0.0023, 0.0069) | 5177 |
| | Log linear | 0.4411 | 0.2985 | (-0.3913, 1.2734) | 5177 |
| | Categorical [Ref. Quartile 1) | | | | |
| | Quartile 2 | -0.0261 | 0.9785 | (-1.9248, 1.8726) | 5181 |
| | Quartile 3 | 0.5091 | 0.601 | (-1.4014, 2.4196) | 5181 |
| | Quartile 4 | 0.9729 | 0.3173 | (-0.9358, 2.8816) | 5181 |
| BC | Linear | 0.0284 | 0.3559 | (-0.032, 0.0888) | 5066 |
| | Log linear | 0.33 | 0.4392 | (-0.507, 1.167) | 5066 |
| | Categorical [Ref. Quartile 1) | | | | |
| | Quartile 2 | 0.3304 | 0.7338 | (-1.5764, 2.2371) | 5068 |
| | Quartile 3 | 1.0504 | 0.2849 | (-0.8766, 2.9775) | 5068 |
| | Quartile 4 | 1.1592 | 0.242 | (-0.7845, 3.1028) | 5068 |
| CO | Linear | 0.0874 | 0.3966 | (-0.115, 0.2898) | 5261 |
| | Log linear | 0.3903 | 0.0604 | (-0.0171, 0.7976) | 5259 |
| | Categorical [Ref. Quartile 1) | | | | |
| | Quartile 2 | 1.2883 | 0.1728 | (-0.5651, 3.1417) | 5262 |
| | Quartile 3 | 1.9096 | 0.042 | (0.0688, 3.7504) | 5262 |
| | Quartile 4 | 1.0558 | 0.2642 | (-0.7993, 2.9108) | 5262 |
| *Diastolic Blood Pressure* | | | | | |
| PM2.5 | Linear | 0.0023 | 0.231 | (-0.0015, 0.006) | 4882 |
| | Log linear | 0.6528 | 0.0584 | (-0.0231, 1.3287) | 4879 |
| | Categorical [Ref. Quartile 1) | | | | |
| | Quartile 2 | -0.3217 | 0.6812 | (-1.8585, 1.2151) | 4878 |
| | Quartile 3 | 1.2127 | 0.1241 | (-0.3337, 2.7591) | 4878 |
| | Quartile 4 | 1.6491 | 0.0365 | (0.1042, 3.194) | 4878 |
| BC | Linear | 0.0271 | 0.282 | (-0.0223, 0.0765) | 4785 |
| | Log linear | 0.5038 | 0.1487 | (-0.1804, 1.1881) | 4784 |
| | Categorical [Ref. Quartile 1) | | | | |
| | Quartile 2 | 0.2138 | 0.7876 | (-1.344, 1.7717) | 4786 |
| | Quartile 3 | 1.2862 | 0.1092 | (-0.2882, 2.8606) | 4786 |
| | Quartile 4 | 1.2893 | 0.1114 | (-0.2987, 2.8773) | 4786 |
| CO | Linear | 0.0833 | 0.3292 | (-0.0842, 0.2507) | 4985 |
| | Log linear | 0.3595 | 0.0365 | (0.0226, 0.6963) | 4981 |
| | Categorical [Ref. Quartile 1) | | | | |
| | Quartile 2 | 0.7423 | 0.3412 | (-0.7878, 2.2724) | 4982 |
| | Quartile 3 | 1.5339 | 0.0479 | (0.0141, 3.0536) | 4982 |
| | Quartile 4 | 1.9873 | 0.0111 | (0.4558, 3.5187) | 4982 |

***Note:*** *All models controlled for gestational age at baseline, BMI, and mother's age, mother's education, household wealth, and season.*

**Table S4b.** Adjusted association between HAP and PP/MAP in India

| | Model Type | Estimate | p-value | 95% CI | AIC |
|---|---|---|---|---|---|
| *Pulse Pressure* | | | | | |
| PM2.5 | Linear | 0 | 0.9965 | (-0.0038, 0.0038) | 4911 |
| | Log linear | -0.2117 | 0.5476 | (-0.9027, 0.4793) | 4911 |
| | Categorical [Ref. Quartile 1) | | | | |
| | Quartile 2 | 0.2956 | 0.7125 | (-1.2789, 1.8701) | 4913 |
| | Quartile 3 | -0.7036 | 0.3835 | (-2.2879, 0.8807) | 4913 |
| | Quartile 4 | -0.6762 | 0.4019 | (-2.259, 0.9066) | 4913 |
| BC | Linear | 0.0013 | 0.9586 | (-0.0486, 0.0512) | 4799 |
| | Log linear | -0.1739 | 0.6216 | (-0.8652, 0.5175) | 4799 |
| | Categorical [Ref. Quartile 1) | | | | |
| | Quartile 2 | 0.1165 | 0.8846 | (-1.46, 1.6931) | 4803 |
| | Quartile 3 | -0.2358 | 0.7715 | (-1.829, 1.3575) | 4803 |
| | Quartile 4 | -0.1302 | 0.8737 | (-1.7372, 1.4769) | 4803 |
| CO | Linear | 0.0042 | 0.9606 | (-0.1612, 0.1695) | 4966 |
| | Log linear | 0.0308 | 0.8562 | (-0.3027, 0.3642) | 4966 |
| | Categorical [Ref. Quartile 1) | | | | |
| | Quartile 2 | 0.546 | 0.4789 | (-0.9671, 2.0591) | 4966 |
| | Quartile 3 | 0.3758 | 0.6236 | (-1.127, 1.8786) | 4966 |
| | Quartile 4 | -0.9315 | 0.2276 | (-2.4459, 0.5829) | 4966 |
| *Mean Arterial Pressure* | | | | | |
| PM2.5 | Linear | 0.0023 | 0.2158 | (-0.0013, 0.0059) | 4837 |
| | Log linear | 0.5822 | 0.0815 | (-0.0731, 1.2375) | 4835 |
| | Categorical [Ref. Quartile 1) | | | | |
| | Quartile 2 | -0.2232 | 0.7691 | (-1.7149, 1.2686) | 4836 |
| | Quartile 3 | 0.9782 | 0.2012 | (-0.5229, 2.4792) | 4836 |
| | Quartile 4 | 1.4237 | 0.0627 | (-0.0759, 2.9233) | 4836 |
| BC | Linear | 0.0275 | 0.259 | (-0.0203, 0.0754) | 4740 |
| | Log linear | 0.4459 | 0.1871 | (-0.217, 1.1088) | 4740 |
| | Categorical [Ref. Quartile 1) | | | | |
| | Quartile 2 | 0.2527 | 0.7424 | (-1.2564, 1.7618) | 4742 |
| | Quartile 3 | 1.2076 | 0.1205 | (-0.3176, 2.7328) | 4742 |
| | Quartile 4 | 1.2459 | 0.1122 | (-0.2924, 2.7843) | 4742 |
| CO | Linear | 0.0847 | 0.3054 | (-0.0774, 0.2468) | 4937 |
| | Log linear | 0.3697 | 0.0263 | (0.0438, 0.6957) | 4933 |
| | Categorical [Ref. Quartile 1) | | | | |
| | Quartile 2 | 0.9243 | 0.2213 | (-0.558, 2.4066) | 4936 |
| | Quartile 3 | 1.6591 | 0.0272 | (0.1869, 3.1313) | 4936 |
| | Quartile 4 | 1.6768 | 0.0268 | (0.1932, 3.1603) | 4936 |

***Note:*** *All models controlled for gestational age at baseline, BMI, and mother's age, mother's education, household wealth, and season.*

**Table S5a.** Adjusted association between HAP and SBP/DBP in Peru

| | Model Type | Estimate | p-value | 95% CI | AIC |
|---|---|---|---|---|---|
| *Systolic Blood Pressure* | | | | | |
| PM2.5 | Linear | 0.0014 | 0.6444 | (-0.0044, 0.0072) | 4384 |
| | Log linear | -0.0235 | 0.9406 | (-0.6412, 0.5943) | 4384 |
| | Categorical [Ref. Quartile 1) | | | | |
| | Quartile 2 | -1.1074 | 0.2131 | (-2.8522, 0.6375) | 4386 |
| | Quartile 3 | -0.4932 | 0.5836 | (-2.2594, 1.273) | 4386 |
| | Quartile 4 | -0.4686 | 0.6009 | (-2.2269, 1.2896) | 4386 |
| BC | Linear | 0.0084 | 0.7706 | (-0.0481, 0.0649) | 3967 |
| | Log linear | -0.2181 | 0.5048 | (-0.86, 0.4238) | 3967 |
| | Categorical [Ref. Quartile 1) | | | | |
| | Quartile 2 | -1.2889 | 0.1745 | (-3.1508, 0.573) | 3967 |
| | Quartile 3 | 0.0841 | 0.9289 | (-1.7666, 1.9348) | 3967 |
| | Quartile 4 | -1.2782 | 0.1729 | (-3.1181, 0.5617) | 3967 |
| CO | Linear | -0.0046 | 0.927 | (-0.103, 0.0938) | 4357 |
| | Log linear | -0.2803 | 0.2227 | (-0.7313, 0.1707) | 4356 |
| | Categorical [Ref. Quartile 1) | | | | |
| | Quartile 2 | -2.0107 | 0.026 | (-3.7805, -0.2408) | 4353 |
| | Quartile 3 | 0.0481 | 0.9574 | (-1.7193, 1.8155) | 4353 |
| | Quartile 4 | -1.5157 | 0.0931 | (-3.2857, 0.2542) | 4353 |
| *Diastolic Blood Pressure* | | | | | |
| PM2.5 | Linear | -0.0038 | 0.1327 | (-0.0088, 0.0012) | 4197 |
| | Log linear | -0.6343 | 0.0193 | (-1.1654, -0.1032) | 4194 |
| | Categorical [Ref. Quartile 1) | | | | |
| | Quartile 2 | -0.9091 | 0.2341 | (-2.4082, 0.5899) | 4196 |
| | Quartile 3 | -2.1263 | 0.0061 | (-3.6437, -0.6089) | 4196 |
| | Quartile 4 | -1.3251 | 0.0855 | (-2.8356, 0.1855) | 4196 |
| BC | Linear | -0.0448 | 0.077 | (-0.0945, 0.0049) | 3821 |
| | Log linear | -0.6638 | 0.0211 | (-1.2274, -0.1002) | 3818 |
| | Categorical [Ref. Quartile 1) | | | | |
| | Quartile 2 | -0.3717 | 0.6557 | (-2.0084, 1.265) | 3820 |
| | Quartile 3 | -1.4124 | 0.0887 | (-3.0392, 0.2144) | 3820 |
| | Quartile 4 | -1.9757 | 0.0167 | (-3.5931, -0.3584) | 3820 |
| CO | Linear | 0.0141 | 0.7423 | (-0.07, 0.0982) | 4161 |
| | Log linear | -0.3945 | 0.0444 | (-0.7792, -0.0099) | 4157 |
| | Categorical [Ref. Quartile 1) | | | | |
| | Quartile 2 | -2.3678 | 0.0022 | (-3.8776, -0.858) | 4155 |
| | Quartile 3 | -1.2191 | 0.1128 | (-2.7268, 0.2886) | 4155 |
| | Quartile 4 | -1.8532 | 0.0162 | (-3.3631, -0.3434) | 4155 |

***Note:*** *All models controlled for gestational age at baseline, BMI, and mother's age, physical activity, time of the BP measurement, household food insecurity, mother's diet diversity and season.*

**Table S5b.** Adjusted association between HAP and PP/MAP in Peru

| | Model Type | Estimate | p-value | 95% CI | AIC |
|---|---|---|---|---|---|
| *Pulse Pressure* | | | | | |
| PM2.5 | Linear | 0.0052 | 0.038 | (0.0003, 0.0101) | 4174 |
| | Log linear | 0.6109 | 0.0219 | (0.0886, 1.1331) | 4173 |
| | Categorical [Ref. Quartile 1) | | | | |
| | Quartile 2 | -0.1983 | 0.7918 | (-1.6726, 1.2761) | 4175 |
| | Quartile 3 | 1.6331 | 0.032 | (0.1408, 3.1254) | 4175 |
| | Quartile 4 | 0.8564 | 0.258 | (-0.6292, 2.342) | 4175 |
| BC | Linear | 0.0532 | 0.0288 | (0.0055, 0.1009) | 3774 |
| | Log linear | 0.4457 | 0.1075 | (-0.0974, 0.9888) | 3776 |
| | Categorical [Ref. Quartile 1) | | | | |
| | Quartile 2 | -0.9171 | 0.2515 | (-2.4866, 0.6524) | 3773 |
| | Quartile 3 | 1.4965 | 0.06 | (-0.0635, 3.0566) | 3773 |
| | Quartile 4 | 0.6975 | 0.3774 | (-0.8534, 2.2484) | 3773 |
| CO | Linear | -0.0187 | 0.6552 | (-0.1008, 0.0634) | 4131 |
| | Log linear | 0.1142 | 0.5517 | (-0.2624, 0.4908) | 4131 |
| | Categorical [Ref. Quartile 1) | | | | |
| | Quartile 2 | 0.3571 | 0.6363 | (-1.1255, 1.8398) | 4132 |
| | Quartile 3 | 1.2671 | 0.0933 | (-0.2134, 2.7477) | 4132 |
| | Quartile 4 | 0.3375 | 0.655 | (-1.1452, 1.8202) | 4132 |
| *Mean Arterial Pressure* | | | | | |
| PM2.5 | Linear | -0.0021 | 0.3851 | (-0.0068, 0.0026) | 4132 |
| | Log linear | -0.4307 | 0.0942 | (-0.9353, 0.0739) | 4130 |
| | Categorical [Ref. Quartile 1) | | | | |
| | Quartile 2 | -0.9752 | 0.1794 | (-2.4, 0.4496) | 4132 |
| | Quartile 3 | -1.5819 | 0.0316 | (-3.0241, -0.1398) | 4132 |
| | Quartile 4 | -1.0396 | 0.1555 | (-2.4753, 0.3961) | 4132 |
| BC | Linear | -0.0271 | 0.2577 | (-0.074, 0.0199) | 3756 |
| | Log linear | -0.5153 | 0.0579 | (-1.0478, 0.0172) | 3754 |
| | Categorical [Ref. Quartile 1) | | | | |
| | Quartile 2 | -0.6774 | 0.3901 | (-2.2247, 0.8698) | 3756 |
| | Quartile 3 | -0.9136 | 0.2438 | (-2.4515, 0.6243) | 3756 |
| | Quartile 4 | -1.7432 | 0.0255 | (-3.2722, -0.2143) | 3756 |
| CO | Linear | 0.0079 | 0.8476 | (-0.0724, 0.0882) | 4103 |
| | Log linear | -0.3565 | 0.0572 | (-0.7238, 0.0109) | 4100 |
| | Categorical [Ref. Quartile 1) | | | | |
| | Quartile 2 | -2.2488 | 0.0023 | (-3.6894, -0.8082) | 4096 |
| | Quartile 3 | -0.7967 | 0.2772 | (-2.2353, 0.6419) | 4096 |
| | Quartile 4 | -1.7407 | 0.018 | (-3.1814, -0.3001) | 4096 |

***Note:*** *All models controlled for gestational age at baseline, BMI, and mother's age, physical activity, time of the BP measurement, household food insecurity, mother's diet diversity and season.*

**Table S6a.** Adjusted association between HAP and SBP/DBP in Rwanda

| | Model Type | Estimate | p-value | 95% CI | AIC |
|---|---|---|---|---|---|
| *Systolic Blood Pressure* | | | | | |
| PM2.5 | Linear | -0.0062 | 0.1083 | (-0.0138, 0.0014) | 5047 |
| | Log linear | -0.3295 | 0.5432 | (-1.3931, 0.7341) | 5049 |
| | Categorical [Ref. Quartile 1) | | | | |
| | Quartile 2 | 0.125 | 0.9068 | (-1.9704, 2.2203) | 5054 |
| | Quartile 3 | -0.1623 | 0.881 | (-2.2906, 1.966) | 5054 |
| | Quartile 4 | -0.0116 | 0.9917 | (-2.1801, 2.1569) | 5054 |
| BC | Linear | 0.0735 | 0.161 | (-0.0294, 0.1764) | 4017 |
| | Log linear | 1.0388 | 0.1603 | (-0.4127, 2.4903) | 4017 |
| | Categorical [Ref. Quartile 1) | | | | |
| | Quartile 2 | -1.899 | 0.1048 | (-4.1953, 0.3973) | 4009 |
| | Quartile 3 | -2.788 | 0.0245 | (-5.2163, -0.3596) | 4009 |
| | Quartile 4 | 1.0233 | 0.4081 | (-1.4046, 3.4511) | 4009 |
| CO | Linear | -0.0626 | 0.4622 | (-0.2298, 0.1045) | 4984 |
| | Log linear | 0.0652 | 0.8124 | (-0.4742, 0.6047) | 4985 |
| | Categorical [Ref. Quartile 1) | | | | |
| | Quartile 2 | 1.2048 | 0.2363 | (-0.791, 3.2006) | 4986 |
| | Quartile 3 | 1.3867 | 0.1704 | (-0.5975, 3.3708) | 4986 |
| | Quartile 4 | 0.2918 | 0.7739 | (-1.7018, 2.2855) | 4986 |
| *Diastolic Blood Pressure* | | | | | |
| PM2.5 | Linear | -0.0077 | 0.0099 | (-0.0136, -0.0019) | 4694 |
| | Log linear | -0.9896 | 0.0184 | (-1.8119, -0.1674) | 4695 |
| | Categorical [Ref. Quartile 1) | | | | |
| | Quartile 2 | 0.019 | 0.9817 | (-1.6035, 1.6415) | 4701 |
| | Quartile 3 | -0.9659 | 0.2502 | (-2.614, 0.6821) | 4701 |
| | Quartile 4 | -1.076 | 0.2088 | (-2.7552, 0.6032) | 4701 |
| BC | Linear | -0.0534 | 0.1786 | (-0.1314, 0.0245) | 3713 |
| | Log linear | -0.8706 | 0.1201 | (-1.9692, 0.228) | 3713 |
| | Categorical [Ref. Quartile 1) | | | | |
| | Quartile 2 | -1.5368 | 0.0851 | (-3.2869, 0.2133) | 3712 |
| | Quartile 3 | -2.3581 | 0.0126 | (-4.2089, -0.5074) | 3712 |
| | Quartile 4 | -1.844 | 0.0508 | (-3.6944, 0.0063) | 3712 |
| CO | Linear | 0.0889 | 0.183 | (-0.042, 0.2198) | 4651 |
| | Log linear | 0.1331 | 0.5365 | (-0.2896, 0.5559) | 4652 |
| | Categorical [Ref. Quartile 1) | | | | |
| | Quartile 2 | 0.5553 | 0.4867 | (-1.0114, 2.1221) | 4656 |
| | Quartile 3 | 0.1787 | 0.8218 | (-1.3789, 1.7363) | 4656 |
| | Quartile 4 | 0.4048 | 0.6117 | (-1.1603, 1.9699) | 4656 |

***Note:*** *All models controlled for gestational age at baseline, BMI, and mother's age, nulliparity, mother's education, physical activity, time of the BP measurement, household food insecurity and smoker at home.*

**Table S6b.** Adjusted association between HAP and PP/MAP in Rwanda

| | Model Type | Estimate | p-value | 95% CI | AIC |
|---|---|---|---|---|---|
| *Pulse Pressure* | | | | | |
| PM2.5 | Linear | 0.0015 | 0.6002 | (-0.0042, 0.0072) | 4655 |
| | Log linear | 0.6602 | 0.1044 | (-0.1371, 1.4574) | 4652 |
| | Categorical [Ref. Quartile 1) | | | | |
| | Quartile 2 | 0.106 | 0.8946 | (-1.4645, 1.6764) | 4656 |
| | Quartile 3 | 0.8036 | 0.3229 | (-0.7915, 2.3988) | 4656 |
| | Quartile 4 | 1.0645 | 0.1989 | (-0.5608, 2.6898) | 4656 |
| BC | Linear | 0.127 | 0.0012 | (0.0501, 0.2038) | 3698 |
| | Log linear | 1.9094 | 6.00E-04 | (0.8268, 2.992) | 3697 |
| | Categorical [Ref. Quartile 1) | | | | |
| | Quartile 2 | -0.3623 | 0.6791 | (-2.0817, 1.3572) | 3693 |
| | Quartile 3 | -0.4299 | 0.6425 | (-2.2482, 1.3885) | 3693 |
| | Quartile 4 | 2.8673 | 0.002 | (1.0493, 4.6853) | 3693 |
| CO | Linear | -0.1515 | 0.0187 | (-0.2777, -0.0253) | 4601 |
| | Log linear | -0.0679 | 0.7444 | (-0.4767, 0.3409) | 4606 |
| | Categorical [Ref. Quartile 1) | | | | |
| | Quartile 2 | 0.6494 | 0.399 | (-0.8617, 2.1605) | 4606 |
| | Quartile 3 | 1.208 | 0.1148 | (-0.2943, 2.7102) | 4606 |
| | Quartile 4 | -0.113 | 0.8832 | (-1.6225, 1.3965) | 4606 |
| *Mean Arterial Pressure* | | | | | |
| PM2.5 | Linear | -0.0072 | 0.0166 | (-0.0131, -0.0013) | 4703 |
| | Log linear | -0.7696 | 0.0686 | (-1.5982, 0.059) | 4705 |
| | Categorical [Ref. Quartile 1) | | | | |
| | Quartile 2 | 0.0543 | 0.948 | (-1.5798, 1.6885) | 4711 |
| | Quartile 3 | -0.6981 | 0.4092 | (-2.3579, 0.9618) | 4711 |
| | Quartile 4 | -0.7212 | 0.4027 | (-2.4124, 0.97) | 4711 |
| BC | Linear | -0.0111 | 0.7828 | (-0.0903, 0.068) | 3730 |
| | Log linear | -0.2341 | 0.6805 | (-1.3506, 0.8823) | 3730 |
| | Categorical [Ref. Quartile 1) | | | | |
| | Quartile 2 | -1.6575 | 0.0668 | (-3.4303, 0.1152) | 3726 |
| | Quartile 3 | -2.5014 | 0.009 | (-4.3761, -0.6267) | 3726 |
| | Quartile 4 | -0.8883 | 0.3523 | (-2.7626, 0.986) | 3726 |
| CO | Linear | 0.0384 | 0.5659 | (-0.0928, 0.1695) | 4653 |
| | Log linear | 0.1105 | 0.6082 | (-0.3126, 0.5336) | 4653 |
| | Categorical [Ref. Quartile 1) | | | | |
| | Quartile 2 | 0.7718 | 0.334 | (-0.7956, 2.3393) | 4656 |
| | Quartile 3 | 0.5814 | 0.4641 | (-0.9769, 2.1396) | 4656 |
| | Quartile 4 | 0.3672 | 0.6454 | (-1.1986, 1.9329) | 4656 |

***Note:*** *All models controlled for gestational age at baseline, BMI, and mother's age, nulliparity, mother's education, physical activity, time of the BP measurement, household food insecurity and smoker at home.*

**Table S7.** Trial-wide association between HAP exposure and blood pressure parameters (without the highest and lowest 2.5% of the exposure samples)

| | $PM_{2.5}$ | | BC | | CO | |
|---|---|---|---|---|---|---|
| | Estimate | 95% CI | Estimate | 95% CI | Estimate | 95% CI |
| ***Systolic Blood Pressure*** | | | | | | |
| Log linear | 0.20 | (-0.24, 0.65) | 0.19 | (-0.54, 0.92) | 0.01 | (-0.33, 0.36) |
| Categorical [Ref. Quartile 1] | | | | | | |
| Quartile 2 | 0.11 | (-0.84, 1.05) | -0.67 | (-1.79, 0.44) | -0.49 | (-2.33, 1.34) |
| Quartile 3 | 0.26 | (-0.69, 1.22) | -0.33 | (-1.90, 1.25) | 0.67 | (-0.49, 1.84) |
| Quartile 4 | 0.50 | (-0.48, 1.48) | 0.13 | (-1.26, 1.52) | -0.37 | (-1.47, 0.74) |
| ***Diastolic Blood Pressure*** | | | | | | |
| Log linear | -0.12 | (-0.94, 0.70) | -0.25 | (-1.28, 0.79) | -0.07 | (-0.59, 0.45) |
| Categorical [Ref. Quartile 1] | | | | | | |
| Quartile 2 | 0.00 | (-0.77, 0.78) | -0.40 | (-1.29, 0.49) | -0.70 | (-2.29, 0.88) |
| Quartile 3 | -0.29 | (-1.78, 1.21) | -0.65 | (-2.31, 1.01) | -0.12 | (-1.43, 1.18) |
| Quartile 4 | 0.05 | (-1.36, 1.46) | -0.74 | (-2.50, 1.02) | -0.16 | (-2.00, 1.69) |
| ***Pulse Pressure*** | | | | | | |
| Log linear | 0.38 | (-0.01, 0.78) | 0.55 | (-0.33, 1.42) | 0.07 | (-0.14, 0.28) |
| Categorical [Ref. Quartile 1] | | | | | | |
| Quartile 2 | 0.12 | (-0.61, 0.85) | -0.31 | (-1.08, 0.46) | 0.14 | (-0.56, 0.85) |
| Quartile 3 | 0.54 | (-0.37, 1.44) | 0.23 | (-0.64, 1.11) | 0.73 | (0.02, 1.44) |
| Quartile 4 | 0.52 | (-0.24, 1.28) | 0.89 | (-0.29, 2.07) | -0.13 | (-0.89, 0.63) |
| ***Mean Arterial Pressure*** | | | | | | |
| Log linear | 0.00 | (-0.69, 0.68) | -0.06 | (-0.92, 0.80) | -0.05 | (-0.51, 0.42) |
| Categorical [Ref. Quartile 1] | | | | | | |
| Quartile 2 | 0.02 | (-0.74, 0.78) | -0.50 | (-1.44, 0.45) | -0.63 | (-2.29, 1.03) |
| Quartile 3 | -0.11 | (-1.31, 1.09) | -0.56 | (-2.14, 1.03) | 0.13 | (-1.09, 1.36) |
| Quartile 4 | 0.06 | (-1.33, 1.45) | -0.43 | (-1.98, 1.13) | -0.22 | (-1.83, 1.40) |

***Notes:***
*1. All models controlled for gestational age at the BP measurement, BMI, and mother's age. Additional covariates are controlled for IRC-specific models (Guatemala: nulliparity, mother's education, physical activity, date of the BP measurement, mother's diet diversity and season; India: mother's education, household wealth, and season; Peru: physical activity, time of the BP measurement, household food insecurity, mother's diet diversity and season; Rwanda: nulliparity, mother's education, physical activity, time of the BP measurement, household food insecurity and smoker at home)*
*2. In log linear models, the coefficients indicate the increase in BP (mmHg) per a one unit increase in the log of exposure.*
*3. Shaded cells are fixed effects, unshaded are random effects, meta-analyses combining results across 4 IRCs*

**Table S8.** Effect modification by gestational age at BP measurement, maternal age, BMI, physical activity, and presence of smoker at home for the association between HAP exposure and BP parameters based on log-linear exposure models in **Guatemala**

| Interaction | Exposure | Estimate | SE | P-value | Estimate | SE | P-value |
|---|---|---|---|---|---|---|---|
| | | | **SBP** | | | **DBP** | |
| Gestational age at baseline BP measurement [Ref. <= Median] | $PM_{2.5}$ | 0.38 | 0.80 | 0.64 | 0.55 | 0.67 | 0.41 |
| | BC | 1.14 | 1.32 | 0.39 | 0.90 | 1.11 | 0.41 |
| | CO | -0.57 | 0.49 | 0.24 | -0.28 | 0.42 | 0.51 |

| | | | | | | | |
|---|---|---|---|---|---|---|---|
| Maternal age [Ref. <= Median] | $PM_{2.5}$ | 0.04 | 0.81 | 0.96 | 0.68 | 0.68 | 0.31 |
| | BC | -0.88 | 1.33 | 0.51 | 0.00 | 1.11 | 1.00 |
| | CO | -0.20 | 0.49 | 0.68 | 0.12 | 0.42 | 0.78 |
| Baseline BMI [Ref. Normal weight] (Coefficients in the order of 'Obese', 'Overweight', and 'Underweight') | $PM_{2.5}$ | 1.85 | 2.08 | 0.38 | 1.44 | 1.75 | 0.41 |
| | | -0.92 | 0.93 | 0.32 | 0.77 | 0.78 | 0.32 |
| | | 4.45 | 5.38 | 0.41 | 6.60 | 4.51 | 0.14 |
| | BC | 2.94 | 3.16 | 0.35 | 1.74 | 2.64 | 0.51 |
| | | -0.81 | 1.45 | 0.58 | 0.96 | 1.22 | 0.43 |
| | | -10.34 | 30.04 | 0.73 | -24.48 | 25.13 | 0.33 |
| | CO | 2.43 | 1.33 | 0.07 | 1.43 | 1.13 | 0.21 |
| | | -1.29 | 0.54 | 0.02 | -0.32 | 0.46 | 0.49 |
| | | 3.29 | 4.56 | 0.47 | 5.40 | 3.89 | 0.17 |
| Baseline physical activity [Ref. <= Median] | $PM_{2.5}$ | -0.13 | 0.80 | 0.88 | 0.51 | 0.67 | 0.45 |
| | BC | 0.16 | 1.35 | 0.91 | -0.39 | 1.13 | 0.73 |
| | CO | -0.10 | 0.49 | 0.84 | 0.12 | 0.42 | 0.78 |
| Smoker present at home [Ref. No/NA] | $PM_{2.5}$ | -2.28 | 1.92 | 0.24 | -0.57 | 1.61 | 0.72 |
| | BC | 0.97 | 4.30 | 0.82 | -2.08 | 3.60 | 0.56 |
| | CO | -1.31 | 1.12 | 0.24 | -0.28 | 0.95 | 0.77 |
| | | | **PP** | | | **MAP** | |
| Gestational age at baseline BP measurement [Ref. <= Median] | $PM_{2.5}$ | -0.17 | 0.56 | 0.76 | 0.50 | 0.67 | 0.46 |
| | BC | 0.24 | 0.95 | 0.80 | 0.98 | 1.09 | 0.37 |
| | CO | -0.30 | 0.34 | 0.39 | -0.38 | 0.41 | 0.36 |
| Maternal age [Ref. <= Median] | $PM_{2.5}$ | -0.64 | 0.56 | 0.25 | 0.47 | 0.67 | 0.48 |
| | BC | -0.88 | 0.96 | 0.36 | -0.29 | 1.10 | 0.79 |
| | CO | -0.32 | 0.34 | 0.35 | 0.01 | 0.41 | 0.98 |
| Baseline BMI [Ref. Normal weight] (Coefficients in the order of 'Obese', 'Overweight', and 'Underweight') | $PM_{2.5}$ | 0.41 | 1.45 | 0.78 | 1.58 | 1.74 | 0.36 |
| | | -1.68 | 0.65 | 0.01 | 0.20 | 0.77 | 0.79 |
| | | -2.15 | 3.75 | 0.57 | 5.88 | 4.48 | 0.19 |
| | BC | 1.20 | 2.27 | 0.60 | 2.14 | 2.61 | 0.41 |
| | | -1.76 | 1.04 | 0.09 | 0.37 | 1.20 | 0.76 |
| | | 14.14 | 21.58 | 0.51 | -19.77 | 24.86 | 0.43 |
| | CO | 1.00 | 0.93 | 0.28 | 1.76 | 1.12 | 0.11 |
| | | -0.96 | 0.38 | 0.01 | -0.64 | 0.46 | 0.16 |
| | | -2.11 | 3.19 | 0.51 | 4.70 | 3.84 | 0.22 |
| Baseline physical activity [Ref. <= Median] | $PM_{2.5}$ | -0.64 | 0.56 | 0.26 | 0.30 | 0.67 | 0.65 |
| | BC | 0.55 | 0.97 | 0.57 | -0.21 | 1.12 | 0.85 |
| | CO | -0.21 | 0.34 | 0.53 | 0.04 | 0.41 | 0.91 |
| Smoker present at home [Ref. No/NA] | $PM_{2.5}$ | -1.71 | 1.34 | 0.20 | -1.14 | 1.60 | 0.48 |
| | BC | 3.05 | 3.10 | 0.32 | -1.06 | 3.56 | 0.77 |
| | CO | -1.02 | 0.78 | 0.19 | -0.62 | 0.94 | 0.51 |

**Table S9.** Effect modification by gestational age at BP measurement, maternal age, BMI, physical activity, and presence of smoker at home for the association between HAP exposure and BP parameters based on log-linear exposure models in **India**

| Interaction | Exposure | Estimate | SE | P-value | Estimate | SE | P-value |
|---|---|---|---|---|---|---|---|
| | | | SBP | | | DBP | |
| Gestational age at baseline BP measurement [Ref. <= Median] | $PM_{2.5}$ | 0.73 | 0.83 | 0.38 | 0.68 | 0.68 | 0.31 |
| | BC | -0.32 | 0.83 | 0.70 | -0.05 | 0.67 | 0.95 |
| | CO | 0.21 | 0.41 | 0.62 | 0.19 | 0.34 | 0.57 |
| Maternal age [Ref. <= Median] | $PM_{2.5}$ | 1.15 | 0.85 | 0.18 | 0.84 | 0.69 | 0.23 |
| | BC | 1.28 | 0.83 | 0.12 | 0.94 | 0.68 | 0.17 |
| | CO | 0.17 | 0.41 | 0.69 | 0.30 | 0.34 | 0.38 |
| Baseline BMI [Ref. Normal weight] (Coefficients in the order of 'Obese', 'Overweight', and 'Underweight') | $PM_{2.5}$ | 7.39 | 3.21 | 0.02 | 2.95 | 2.60 | 0.26 |
| | | -0.48 | 1.96 | 0.81 | -3.26 | 1.59 | 0.04 |
| | | 0.15 | 0.89 | 0.87 | 0.21 | 0.72 | 0.77 |
| | BC | 14.23 | 5.16 | 0.01 | 5.87 | 4.22 | 0.16 |
| | | 1.26 | 1.71 | 0.46 | -1.96 | 1.40 | 0.16 |
| | | -0.08 | 0.89 | 0.93 | 0.16 | 0.73 | 0.82 |
| | CO | 1.45 | 1.68 | 0.39 | 0.63 | 1.39 | 0.65 |
| | | 1.37 | 0.94 | 0.15 | 0.98 | 0.78 | 0.21 |
| | | -0.27 | 0.43 | 0.53 | 0.05 | 0.36 | 0.89 |
| Baseline physical activity [Ref. <= Median] | $PM_{2.5}$ | -0.01 | 0.83 | 0.99 | 0.00 | 0.68 | 1.00 |
| | BC | 0.98 | 0.83 | 0.24 | 0.13 | 0.68 | 0.84 |
| | CO | 0.52 | 0.42 | 0.21 | 0.24 | 0.34 | 0.49 |
| Smoker present at home [Ref. No/NA] | $PM_{2.5}$ | -0.84 | 0.89 | 0.35 | -0.48 | 0.73 | 0.50 |
| | BC | -0.33 | 0.90 | 0.71 | -0.16 | 0.74 | 0.83 |
| | CO | 0.57 | 0.44 | 0.19 | 0.64 | 0.36 | 0.08 |
| | | | PP | | | MAP | |
| Gestational age at baseline BP measurement [Ref. <= Median] | $PM_{2.5}$ | 0.05 | 0.69 | 0.95 | 0.70 | 0.65 | 0.29 |
| | BC | -0.27 | 0.68 | 0.69 | -0.14 | 0.65 | 0.83 |
| | CO | 0.01 | 0.34 | 0.97 | 0.20 | 0.33 | 0.55 |
| Maternal age [Ref. <= Median] | $PM_{2.5}$ | 0.32 | 0.71 | 0.66 | 0.94 | 0.67 | 0.16 |
| | BC | 0.34 | 0.69 | 0.62 | 1.05 | 0.66 | 0.11 |
| | CO | -0.14 | 0.34 | 0.69 | 0.26 | 0.33 | 0.44 |
| Baseline BMI [Ref. Normal weight] (Coefficients in the order of 'Obese', 'Overweight', and 'Underweight') | $PM_{2.5}$ | 4.44 | 2.66 | 0.10 | 4.43 | 2.52 | 0.08 |
| | | 2.78 | 1.62 | 0.09 | -2.34 | 1.54 | 0.13 |
| | | -0.06 | 0.74 | 0.93 | 0.19 | 0.70 | 0.79 |
| | BC | 8.36 | 4.26 | 0.05 | 8.66 | 4.09 | 0.03 |
| | | 3.21 | 1.41 | 0.02 | -0.88 | 1.35 | 0.51 |
| | | -0.24 | 0.73 | 0.74 | 0.08 | 0.70 | 0.91 |
| | CO | 0.82 | 1.37 | 0.55 | 0.91 | 1.34 | 0.50 |
| | | 0.38 | 0.77 | 0.62 | 1.11 | 0.75 | 0.14 |
| | | -0.32 | 0.35 | 0.36 | -0.06 | 0.35 | 0.87 |
| Baseline physical activity [Ref. <= Median] | $PM_{2.5}$ | -0.01 | 0.69 | 0.99 | 0.00 | 0.65 | 1.00 |
| | BC | 0.85 | 0.69 | 0.22 | 0.42 | 0.66 | 0.53 |

| | | | | | | | |
|---|---|---|---|---|---|---|---|
| | CO | 0.28 | 0.34 | 0.40 | 0.33 | 0.33 | 0.32 |
| Smoker present at home [Ref. No/NA] | $PM_{2.5}$ | -0.36 | 0.74 | 0.63 | -0.60 | 0.70 | 0.39 |
| | BC | -0.17 | 0.75 | 0.81 | -0.22 | 0.71 | 0.76 |
| | CO | -0.07 | 0.36 | 0.85 | 0.62 | 0.35 | 0.08 |

**Table S10.** Effect modification by gestational age at BP measurement, maternal age, BMI, physical activity, and presence of smoker at home for the association between HAP exposure and BP parameters based on log-linear exposure models in **Peru**

| Interaction | Exposure | Estimate | SE | P-value | Estimate | SE | P-value |
|---|---|---|---|---|---|---|---|
| | | | **SBP** | | | **DBP** | |
| Gestational age at baseline BP measurement [Ref. <= Median] | $PM_{2.5}$ | 0.80 | 0.63 | 0.21 | -0.52 | 0.54 | 0.34 |
| | BC | 0.15 | 0.65 | 0.82 | -0.70 | 0.57 | 0.22 |
| | CO | -0.08 | 0.46 | 0.87 | 0.32 | 0.39 | 0.42 |
| Maternal age [Ref. <= Median] | $PM_{2.5}$ | -0.52 | 0.63 | 0.41 | -1.44 | 0.54 | 0.01 |
| | BC | -0.27 | 0.65 | 0.68 | -1.38 | 0.57 | 0.02 |
| | CO | -0.94 | 0.45 | 0.04 | -0.77 | 0.39 | 0.05 |
| Baseline BMI [Ref. Normal weight] (Coefficients in the order of 'Obese', 'Overweight', and 'Underweight') | $PM_{2.5}$ | 2.36 | 0.99 | 0.02 | 0.81 | 0.85 | 0.34 |
| | | 1.05 | 0.67 | 0.12 | 0.14 | 0.58 | 0.81 |
| | | NA | NA | NA | NA | NA | NA |
| | BC | 2.26 | 1.05 | 0.03 | 0.88 | 0.93 | 0.34 |
| | | 0.99 | 0.70 | 0.16 | 0.26 | 0.61 | 0.67 |
| | | NA | NA | NA | NA | NA | NA |
| | CO | 1.15 | 0.72 | 0.11 | 0.78 | 0.61 | 0.21 |
| | | -0.34 | 0.49 | 0.48 | 0.12 | 0.42 | 0.78 |
| | | NA | NA | NA | NA | NA | NA |
| Baseline physical activity [Ref. <= Median] | $PM_{2.5}$ | 0.10 | 0.63 | 0.87 | -0.59 | 0.54 | 0.27 |
| | BC | -0.30 | 0.65 | 0.64 | -0.84 | 0.57 | 0.14 |
| | CO | -0.16 | 0.45 | 0.72 | -0.60 | 0.39 | 0.12 |
| Smoker present at home [Ref. No/NA] | $PM_{2.5}$ | 0.87 | 3.87 | 0.82 | -1.23 | 3.33 | 0.71 |
| | BC | -2.56 | 4.75 | 0.59 | -4.95 | 4.17 | 0.24 |
| | CO | 2.84 | 6.41 | 0.66 | 3.04 | 5.46 | 0.58 |
| | | | **PP** | | | **MAP** | |
| Gestational age at baseline BP measurement [Ref. <= Median] | $PM_{2.5}$ | 1.32 | 0.53 | 0.01 | -0.08 | 0.51 | 0.87 |
| | BC | 0.85 | 0.55 | 0.12 | -0.42 | 0.54 | 0.44 |
| | CO | -0.39 | 0.38 | 0.31 | 0.19 | 0.38 | 0.62 |
| Maternal age [Ref. <= Median] | $PM_{2.5}$ | 0.92 | 0.53 | 0.08 | -1.13 | 0.51 | 0.03 |
| | BC | 1.11 | 0.55 | 0.04 | -1.01 | 0.54 | 0.06 |
| | CO | -0.17 | 0.38 | 0.65 | -0.83 | 0.37 | 0.03 |
| Baseline BMI [Ref. Normal weight] (Coefficients in the order of 'Obese', 'Overweight', and 'Underweight') | $PM_{2.5}$ | 1.55 | 0.83 | 0.06 | 1.32 | 0.81 | 0.10 |
| | | 0.91 | 0.57 | 0.11 | 0.44 | 0.55 | 0.42 |
| | | NA | NA | NA | NA | NA | NA |
| | BC | 1.38 | 0.89 | 0.12 | 1.34 | 0.87 | 0.13 |
| | | 0.73 | 0.59 | 0.22 | 0.50 | 0.58 | 0.39 |

| | | | | | | | |
|---|---|---|---|---|---|---|---|
| | | NA | NA | NA | NA | NA | NA |
| | CO | 0.37 | 0.60 | 0.53 | 0.90 | 0.59 | 0.12 |
| | | -0.46 | 0.40 | 0.26 | -0.04 | 0.40 | 0.93 |
| | | NA | NA | NA | NA | NA | NA |
| Baseline physical activity [Ref. <= Median] | $PM_{2.5}$ | 0.70 | 0.53 | 0.19 | -0.36 | 0.51 | 0.48 |
| | BC | 0.53 | 0.55 | 0.33 | -0.66 | 0.54 | 0.22 |
| | CO | 0.44 | 0.38 | 0.25 | -0.45 | 0.37 | 0.22 |
| Smoker present at home [Ref. No/NA] | $PM_{2.5}$ | 2.09 | 3.27 | 0.52 | -0.53 | 3.16 | 0.87 |
| | BC | 2.38 | 4.02 | 0.55 | -4.15 | 3.94 | 0.29 |
| | CO | -0.20 | 5.35 | 0.97 | 2.97 | 5.22 | 0.57 |

**Table S11.** Effect modification by gestational age at BP measurement, maternal age, BMI, physical activity, and presence of smoker at home for the association between HAP exposure and BP parameters based on log-linear exposure models in **Rwanda**

| Interaction | Exposure | Estimate | SE | P-value | Estimate | SE | P-value |
|---|---|---|---|---|---|---|---|
| | | | **SBP** | | | **DBP** | |
| Gestational age at baseline BP measurement [Ref. <= Median] | $PM_{2.5}$ | -0.14 | 0.99 | 0.88 | 0.82 | 0.76 | 0.28 |
| | BC | -0.48 | 1.33 | 0.72 | 0.28 | 1.01 | 0.78 |
| | CO | 0.35 | 0.55 | 0.52 | 0.09 | 0.43 | 0.84 |
| Maternal age [Ref. <= Median] | $PM_{2.5}$ | -1.02 | 1.00 | 0.31 | -0.30 | 0.77 | 0.69 |
| | BC | -0.88 | 1.35 | 0.51 | -0.66 | 1.02 | 0.52 |
| | CO | -0.03 | 0.55 | 0.96 | -0.78 | 0.43 | 0.07 |
| Baseline BMI [Ref. Normal weight] (Coefficients in the order of 'Obese', 'Overweight', and 'Underweight') | $PM_{2.5}$ | 2.88 | 2.31 | 0.21 | 2.01 | 1.79 | 0.26 |
| | | -0.24 | 1.22 | 0.85 | -0.47 | 0.95 | 0.62 |
| | | -3.06 | 4.08 | 0.45 | 0.90 | 3.16 | 0.78 |
| | BC | 1.83 | 3.03 | 0.55 | 1.88 | 2.29 | 0.41 |
| | | -0.86 | 1.68 | 0.61 | -1.10 | 1.27 | 0.39 |
| | | 0.15 | 5.35 | 0.98 | -0.68 | 4.04 | 0.87 |
| | CO | -0.66 | 1.02 | 0.52 | -0.52 | 0.80 | 0.51 |
| | | 0.90 | 0.69 | 0.19 | 0.67 | 0.54 | 0.21 |
| | | -1.96 | 2.18 | 0.37 | -0.49 | 1.71 | 0.77 |
| Baseline physical activity [Ref. <= Median] | $PM_{2.5}$ | -0.05 | 1.05 | 0.96 | -0.48 | 0.81 | 0.56 |
| | BC | -1.29 | 1.42 | 0.36 | -1.58 | 1.07 | 0.14 |
| | CO | -0.60 | 0.55 | 0.28 | -0.32 | 0.43 | 0.46 |
| Smoker present at home [Ref. No/NA] | $PM_{2.5}$ | 1.17 | 2.36 | 0.62 | 0.30 | 1.83 | 0.87 |
| | BC | 0.11 | 2.87 | 0.97 | 0.64 | 2.17 | 0.77 |
| | CO | -0.80 | 1.83 | 0.66 | -0.62 | 1.43 | 0.66 |
| | | | **PP** | | | **MAP** | |
| Gestational age at baseline BP measurement [Ref. <= Median] | $PM_{2.5}$ | -0.96 | 0.74 | 0.19 | 0.50 | 0.77 | 0.52 |
| | BC | -0.76 | 0.99 | 0.44 | 0.03 | 1.02 | 0.98 |
| | CO | 0.27 | 0.41 | 0.52 | 0.18 | 0.43 | 0.68 |
| Maternal age [Ref. <= Median] | $PM_{2.5}$ | -0.71 | 0.75 | 0.34 | -0.54 | 0.78 | 0.49 |
| | BC | -0.22 | 1.00 | 0.83 | -0.73 | 1.03 | 0.48 |
| | CO | 0.75 | 0.42 | 0.07 | -0.53 | 0.43 | 0.22 |

| | | | | | | | |
|---|---|---|---|---|---|---|---|
| Baseline BMI [Ref. Normal weight] (Coefficients in the order of 'Obese', 'Overweight', and 'Underweight') | $PM_{2.5}$ | 0.87 | 1.71 | 0.61 | 2.30 | 1.81 | 0.20 |
| | | 0.23 | 0.91 | 0.80 | -0.39 | 0.96 | 0.68 |
| | | -3.95 | 3.02 | 0.19 | -0.42 | 3.19 | 0.90 |
| | BC | -0.05 | 2.24 | 0.98 | 1.86 | 2.33 | 0.42 |
| | | 0.23 | 1.24 | 0.85 | -1.02 | 1.29 | 0.43 |
| | | 0.83 | 3.96 | 0.83 | -0.41 | 4.12 | 0.92 |
| | CO | -0.14 | 0.77 | 0.86 | -0.57 | 0.80 | 0.48 |
| | | 0.23 | 0.52 | 0.65 | 0.75 | 0.54 | 0.17 |
| | | -1.47 | 1.64 | 0.37 | -0.98 | 1.71 | 0.57 |
| Baseline physical activity [Ref. <= Median] | $PM_{2.5}$ | 0.43 | 0.79 | 0.59 | -0.33 | 0.82 | 0.68 |
| | BC | 0.29 | 1.06 | 0.78 | -1.48 | 1.09 | 0.17 |
| | CO | -0.29 | 0.42 | 0.49 | -0.41 | 0.43 | 0.34 |
| Smoker present at home [Ref. No/NA] | $PM_{2.5}$ | 0.87 | 1.77 | 0.62 | 0.59 | 1.84 | 0.75 |
| | BC | -0.53 | 2.14 | 0.81 | 0.46 | 2.21 | 0.83 |
| | CO | -0.18 | 1.38 | 0.90 | -0.68 | 1.43 | 0.64 |

**Table S12**. Summary of SBP, DBP and selected baseline characteristics in population with and without missing exposures to $PM_{2.5}$, BC, and CO

| | **$PM_{2.5}$** | | **BC** | | **CO** | |
|---|---|---|---|---|---|---|
| | Missing | Non-missing | Missing | Non-missing | Missing | Non-missing |
| SBP (mmHg), Mean (SD) | 103.6 (8.9)* | 105.0 (9.8)* | 104.6 (10.0) | 104.9 (9.6) | 103.5 (9.6)* | 105.0 (9.7)* |
| DBP (mmHg), Mean (SD) | 60.3 (7.6) | 60.8 (7.8) | 60.6 (8.0) | 60.8 (7.8) | 60.1 (7.7) | 60.8 (7.8) |
| Maternal age (years), Mean (SD) | 25.2 (4.4) | 25.4 (4.5) | 25.6 (4.4) | 25.3 (4.5) | 25.4 (4.3) | 25.4 (4.5) |
| BMI (kg/m2), Mean (SD) | 23.3 (4.2) | 23.2 (4.0) | 23.6 (4.2)* | 23.1 (4.0)* | 24.1 (4.0)* | 23.1 (4.1)* |
| Nulliparous, N (%) | | | | | | |
| Yes | 134 (36%) | 1094 (29%) | 219 (34%) | 1009 (40%) | 126 (40%) | 1102 (38%) |
| No | 238 (64%) | 1718 (61%) | 434 (66%) | 1522 (60%) | 191 (60%) | 1765 (62%) |
| Missing | 0 | 6 (<1%) | 1 (<1%) | 5 (<1%) | 1 (<1%) | 5 (<1%) |
| Mother's Education, N (%) | | | * | * | | |
| No formal education or Primary school incomplete | 127 (34%) | 912 (32%) | 234 (36%) | 805 (32%) | 63 (20%) | 976 (34%) |
| Primary school complete or Secondary school incomplete | 122 (33%) | 967 (34%) | 229 (35%) | 860 (34%) | 109 (34%) | 980 (34%) |
| Secondary school complete or Vocational or Some college or university | 122 (33%) | 939 (33%) | 190 (29%) | 871 (34%) | 146 (46%) | 915 (32%) |
| Missing | 1 (<1%) | 0 | 1 (<1%) | 0 | 0 | 1 (<1%) |

*Note:*
*1. * indicates statistical significance at $\alpha = 0.05$. T-tests were performed for continuous variable and chi-square tests were performed for categorical variable.*
*2. Summary based on the 3190 pregnant women included in the analysis.*

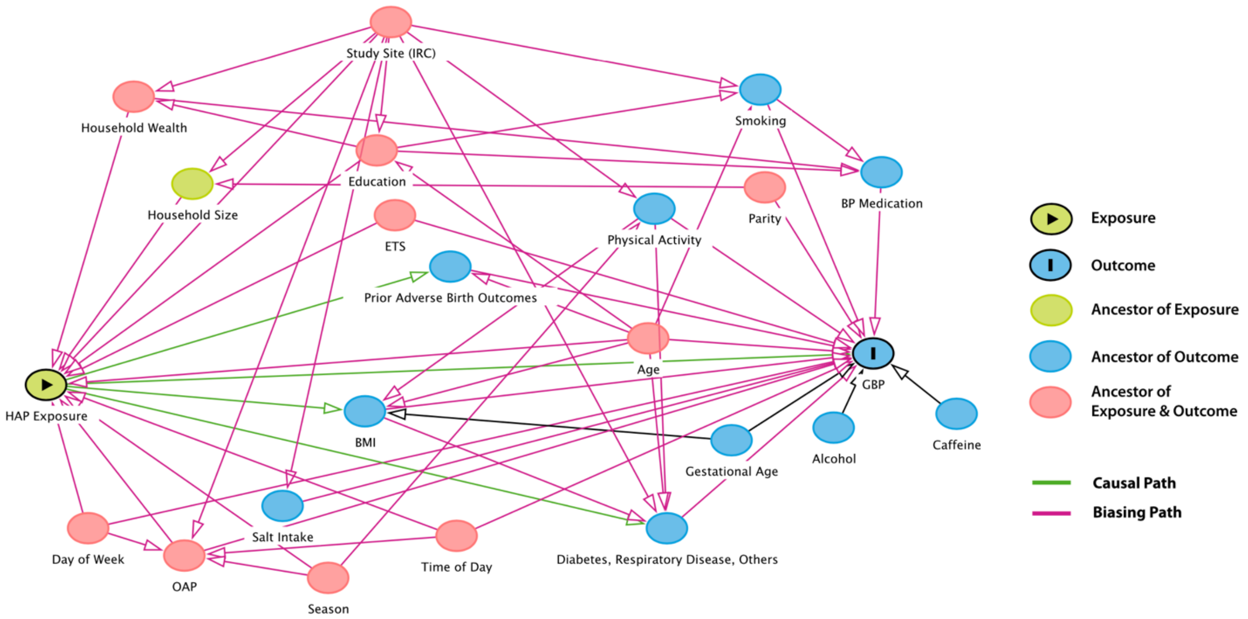


**Figure S1**. Directed Acyclic Graph (DAG) for the association between household air pollution (HAP) exposure from solid fuels and gestation blood pressure (GBP)

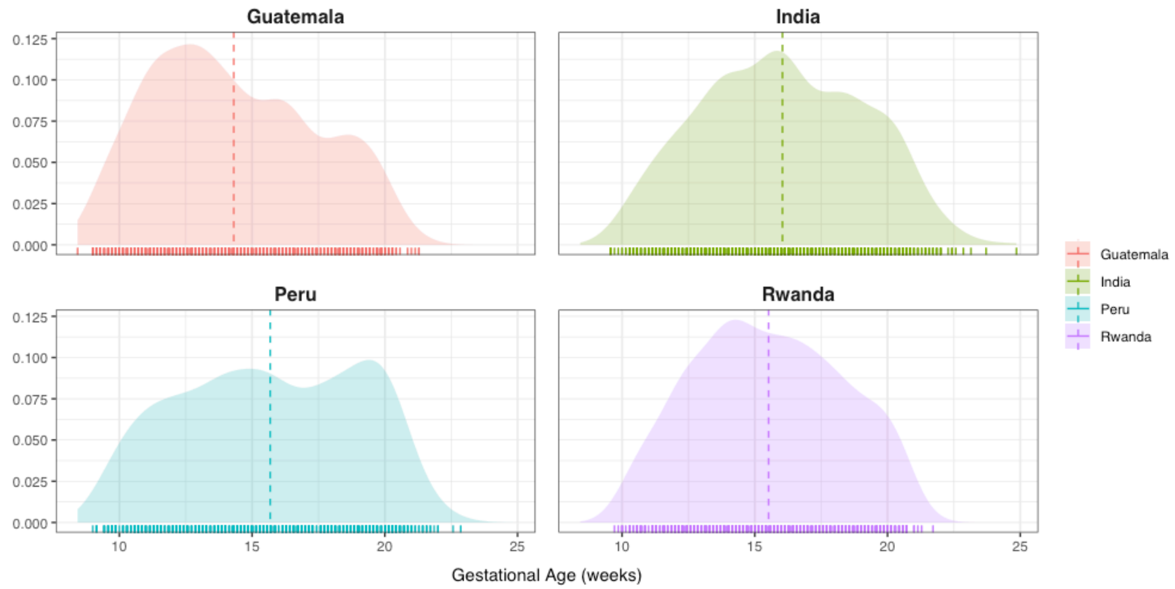


**Figure S2.** Distributions of gestational age at baseline. Solid lines along the x-axes are individual data points. Dashed lines are mean values.

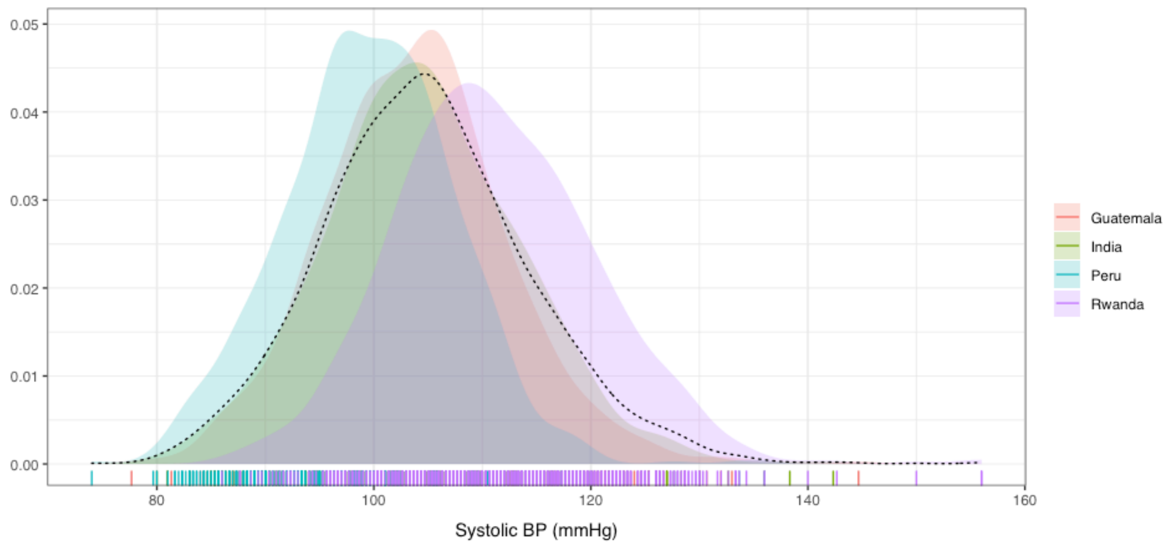


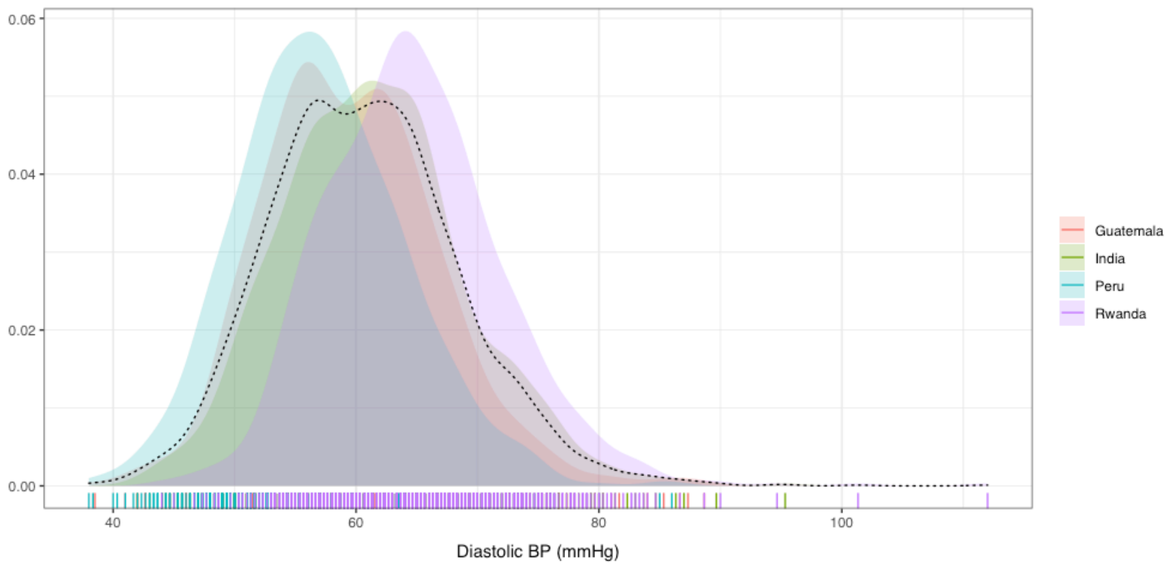


**Figure S3.** Distributions of systolic (top) and diastolic (bottom) blood pressure at baseline. Filled density plots are individual IRC distributions; the dotted line with no fill is the HAPIN-wide distribution. Individual data points are shown as bars along the x-axes.

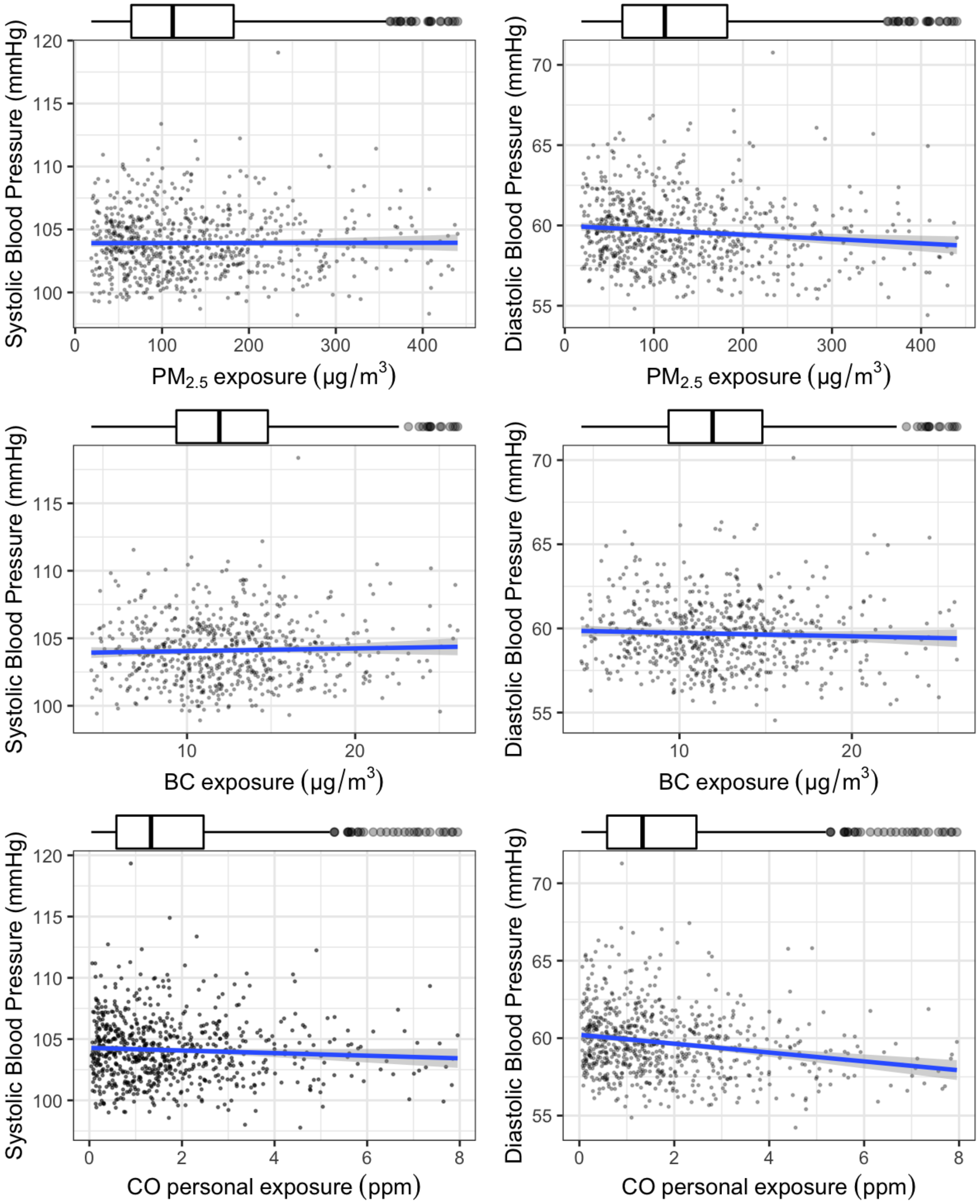


**Figure S4.** HAP-GBP association (blue line) and 95% confidence intervals (shade) in **Guatemala** generated from generalized additive models (GAMs) controlling for gestational age at the BP measurement, BMI, and mother's age, nulliparity, mother's education, physical activity, date of the BP measurement (weekday vs. weekend), mother's diet diversity and season (dry vs. rainy). All plots are showing 95% (after removing the highest and lowest 2.5%) of exposure the data for each household air pollutant.

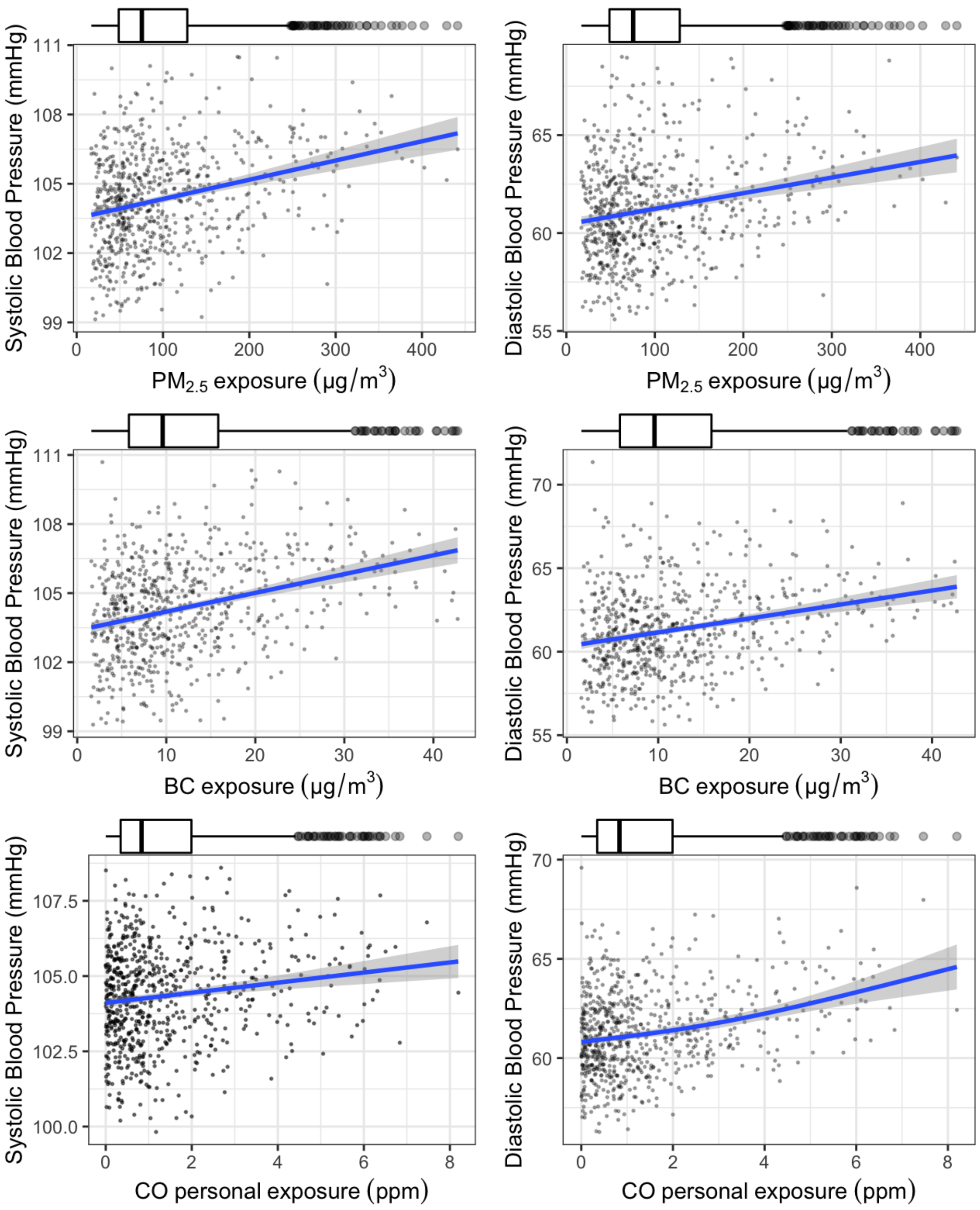


**Figure S5.** HAP-GBP association (blue line) and 95% confidence intervals (shade) in **India** generated from generalized additive models (GAMs) controlling for gestational age at baseline, BMI, mother's age, mother's education, household wealth, and season (winter, summer, and monsoon). All plots are showing 95% (after removing the highest and lowest 2.5%) of exposure the data for each household air pollutant.

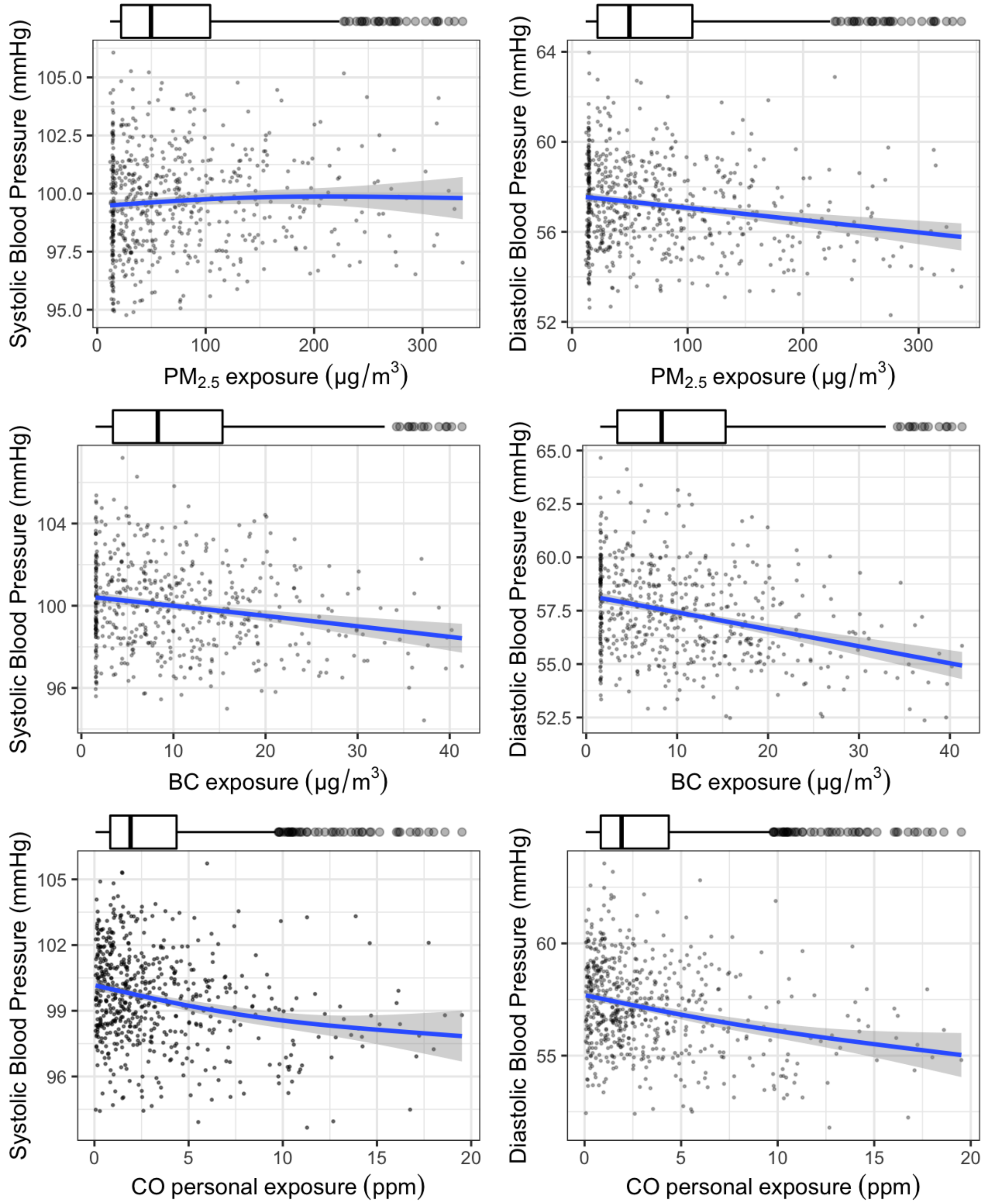


**Figure S6.** HAP-GBP association (blue line) and 95% confidence intervals (shade) in **Peru** generated from generalized additive models (GAMs) controlling for gestational age at baseline, BMI, and mother's age, physical activity, time of the BP measurement (morning vs. afternoon), household food insecurity, mother's diet diversity and season (dry vs. rainy). All plots are showing 95% (after removing the highest and lowest 2.5%) of exposure the data for each household air pollutant.

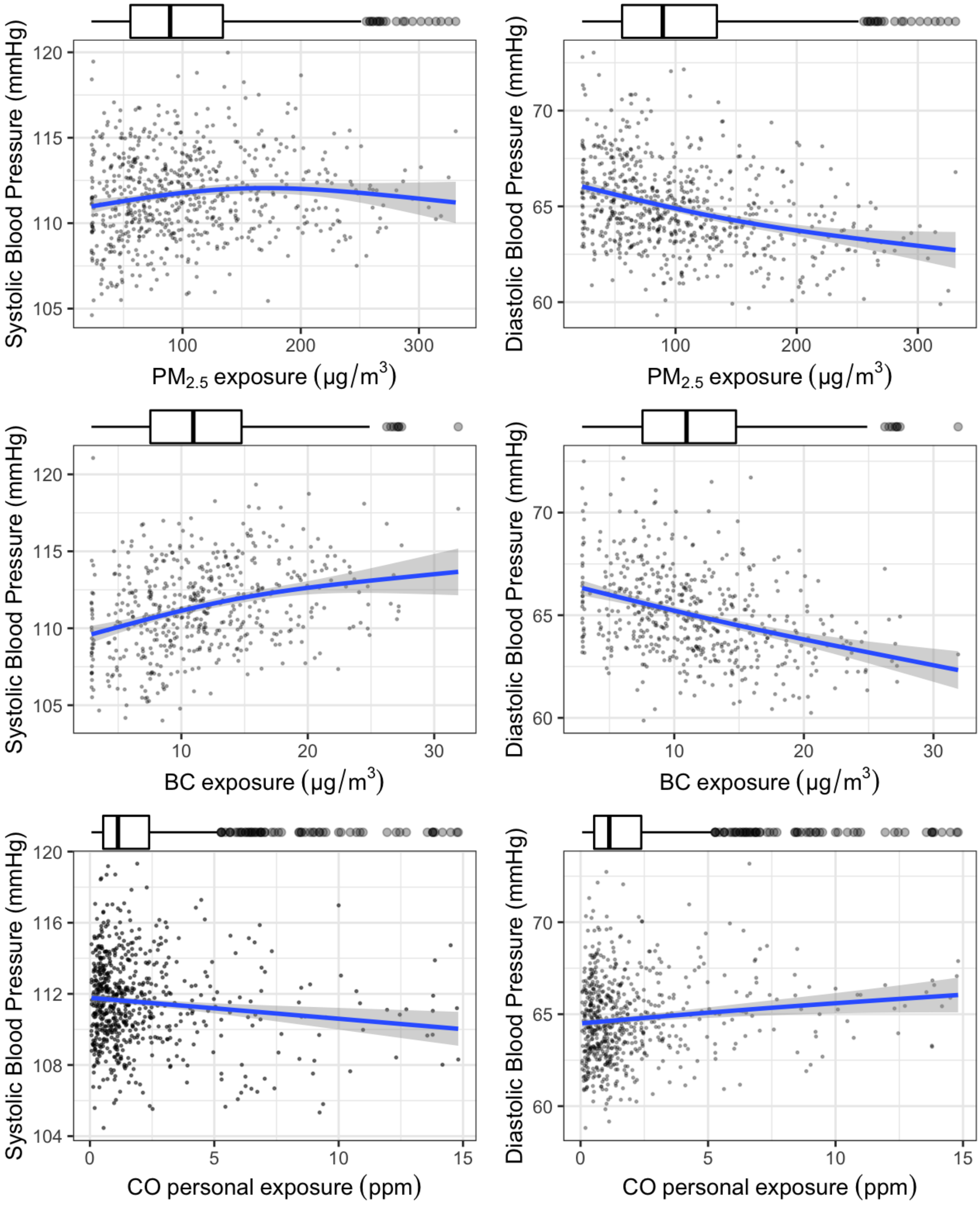


**Figure S7.** HAP-GBP association (blue line) and 95% confidence intervals (shade) in **Rwanda** generated from generalized additive models (GAMs) controlling for gestational age at baseline, BMI, and mother's age, nulliparity, mother's education, physical activity, time of the BP measurement (morning vs. afternoon), household food insecurity and smoker at home. All plots are showing 95% (after removing the highest and lowest 2.5%) of exposure the data for each household air pollutant.
